# Pre-pandemic mental health and brain characteristics predict adolescent stress and emotions during the COVID-19 pandemic

**DOI:** 10.1101/2025.05.12.25327434

**Authors:** Matthew Risner, Linfeng Hu, Catherine Stamoulis

## Abstract

**Background:** The COVID-19 pandemic had profound effects on developing adolescents that, to date, remain incompletely understood. Youth with preexisting mental health problems and associated brain alterations were at increased risk for higher stress and poor mental health. This study investigated impacts of adolescent pre-pandemic mental health problems and their neural correlates on stress, negative emotions and poor mental health during the first ∼15 months of the COVID-19 pandemic.

**Methods:** N=2,641 adolescents (median age=12.0 years) from the Adolescent Brain Cognitive Development (ABCD) cohort, with pre-pandemic data on anxiety, depression, and behavioral (attention, aggression, social withdrawal, internalizing, externalizing) problems, longitudinal survey data on mental health, stress and emotions during the first ∼15 months following the outbreak, structural MRI, and resting-state fMRI. Data were analyzed using mixed effects mediation and moderation models.

**Results:** Preexisting mental health and behavioral problems predicted higher stress, negative affect and negative emotions (β=0.09-0.21, CI=[0.03,0.32]), and lower positive affect (β=-0.21 to -0.09, CI=[-0.31,-0.01]) during the first ∼6 months of the outbreak. Pre-pandemic structural characteristics of brain regions supporting social function and emotional processing (insula, superior temporal gyrus, orbitofrontal cortex, and the cerebellum) mediated some of these relationships (β=0.10-0.15, CI=[0.01,0.24]). Pre-pandemic brain circuit organization moderated (attenuated) relationships between preexisting mental health and pandemic stress and negative emotions (β=-0.17 to -0.06, CI=[-0.27,-0.01]).

**Conclusions:** Preexisting mental health problems and their structural brain correlates were risk factors for youth stress and emotions during the early months of the outbreak. In addition, the organization of some brain circuits was protective and attenuated the effects of preexisting mental health issues on youth responses to the pandemic’s stressors.

## 1. INTRODUCTION

Environmental and experiential factors during adolescence contribute to profound and complex biological changes^1-4^ with important implications for physical and mental health^5-8^. Negative experiences and environmental stressors during this sensitive period significantly increase the risk of neural miswiring^9-13^, which, in turn, can lead to mental health issues and the emergence of neuropsychiatric disorders^7,14-16^.

Recent studies have specifically sought to elucidate how environmental stressors associated with the COVID-19 pandemic impacted adolescent mental health and brain development. They have shown that prolonged stress, social isolation, school-related disruptions, decreased physical activity, and increased media use, had extensive negative effects on brain development and mental health^17-18^, especially in girls and youth from racio-ethnic minorities^19-20^. Preexisting mental health problems that are common in adolescents–such as anxiety, depression, internalizing and externalizing problems–increased vulnerability to these stressors^21-23^. A study on ∼5,000 youth from the Adolescent Brain Cognitive Development (ABCD) cohort^24^ reported exacerbated depressive symptoms, attention problems, and aggressive behaviors in youth with preexisting mental health problems^25^. Another study in ∼8,000 youth from the same cohort showed that history of adverse childhood experiences was a significant risk factor for poor mental health, increased stress, and fear during the pandemic, especially in those with preexisting internalizing problems^19^. Recent work on brain markers as risk or protective factors has shown that stronger and more resilient neural circuits before the outbreak were protective, whereas weaker and more fragile ones increased risk of stress and negative emotions^26^.

The relationship between youth pre-pandemic mental health/behavioral problems and their neural substrates, and their combined (mediating or moderating) impacts on mental health, emotional responses and stress during the pandemic are incompletely understood. A prior study in young adults showed that functional brain connectivity prior to the pandemic predicted pandemic-related anxiety^27^. Another study identified psychological resilience as a mediator of the association between functional connectivity of the Default Mode Network (DMN) and the inferior temporal gyrus and traumatic stress during the pandemic^28^. A recent study on ∼2,600 adolescents from the ABCD cohort reported predictive relationships between pre-pandemic topological characteristics of the underdeveloped salience network and the prefrontal cortex and stress and negative affect during the pandemic^26^. However, this work did not examine the effects of pre-pandemic mental health and behavioral problems on developing brain circuits, which may have, in turn, have increased risk for poor outcomes during the pandemic. Collectively, prior studies have examined pre-pandemic mental health problems and brain characteristics separately, but none has examined their relationships, interactions, and combined impacts on youth responses to the pandemic’s stressors.

This study aimed to address this important gap in knowledge, and elucidate the impact of pre-pandemic mental health on youth responses to the pandemic stressors, directly and indirectly through its effects on the brain. It investigated direct, mediating, moderating and interacting relationships between preexisting mental health issues, the topological organization and morphometric properties of brain circuits and their constituent regions, and pandemic outcomes, including overall mental health, stress, and emotional responses during the first ∼15 months following the outbreak. In 2,641 adolescents from the ABCD cohort with neuroimaging data and assessments of common mental health and behavioral problems, and longitudinal survey data collected during the pandemic, the study tested two hypotheses: a) pre-pandemic anxiety, depression and/or behavioral problems (attention problems, aggressive, internalizing and/or externalizing behaviors, preference for solitude, and/or social withdrawal) predicted worse mental health, higher stress and negative emotions during the pandemic directly and through the mediating effects of brain circuits adversely modulated by these problems; b) the organization of brain circuits and their structural characteristics moderated the relationships between pre-pandemic mental health issues and pandemic responses. Finally, it also hypothesized that these relationships were also affected by protective factors, specifically parental engagement and youth spirituality/religiosity. Prior studies have shown that these factors had positive effects on youth mental health during the pandemic^25,29^.

## 2. METHODS

This study was approved by the Institutional Review Board. All analyzed data were from the ABCD release 4.0 and are publicly available through the National Institute of Mental Health Data Archive (NDA).

### 2.1 Participants

Youth from the 2-year follow-up ABCD cohort were studied (n = 2,641; median age at fMRI scan = 12.0 years, interquartile range (IQR) = 1.1 years). The sample included youth with common mental health problems, such as depressive symptoms and anxiety, but excluded psychiatric and neurodevelopmental disorders (including schizophrenia, psychotic disorders, Autism Spectrum Disorders, Attention-Deficit/Hyperactivity Disorder), all of which adversely modulate developing brain structures and circuits in disorder-specific ways and deserve dedicated studies. The analytic sample consisted of two overlapping cohorts to facilitate sensitivity and replication analyses: a) *n = 1,414* (cohort A) with fMRI scans collected ≤9 months prior to any pandemic-related survey. This cutoff was used to minimize potential developmental brain changes from scan to survey (median scan-to-survey time was 3-7 months). In this cohort, 460 participants had fMRI scans collected during the pandemic; b) *n = 2,174* (cohort B) with fMRI data collected prior to March 11, 2020, when the World Health Organization declared COVID-19 a pandemic^30^ (median scan-to-survey time was 10-22 months). Appropriate adjustments were included in statistical analyses to account for pandemic-related effects on the brain (in cohort A) and pubertal changes (in cohort B).

Youth responses from seven pandemic surveys were analyzed. Not all youth had data across surveys. Thus, seven partially overlapping sub-cohorts were studied with sample sizes from n = 802 (May 2020) to n = 218 (May 2021) in cohort A and n = 1451 (May 2020) to n = 1135 (May 2021) in cohort B. Youth demographic characteristics in cohorts A and B are summarized in Table 1. Boys and girls were approximately equally distributed in the sample. Less than 50% were racio-ethnic minorities and ∼20% were Hispanic. Median BMI was 19.2 kg/m^2^ (IQR = 5.4). Over 30% were in mid-puberty (888 (33.6%)), ∼20% in early puberty (560 (21.2%)), and ∼20% in late puberty (555 (21.0%)).

**Table 1.** Demographic data for the overlapping study cohorts (A and B) and those in cohort B not included in A. The ‘other’ non-Hispanic race-ethnicity category included participants from groups underrepresented in these cohorts (and more broadly the ABCD study): Filipino, Vietnamese, Alaska Native, American Indian, Asian Indian, Chinese, Guamanian, Hawaiian, Japanese, Korean, Native Samoan, other Pacific Islander, other Asian, participants who selected more than 1 racial groups, and those who selected ‘other race’ when reporting demographics.

|  |  | **N = 1414**<br>**(Cohort A)** | **N = 2174**<br>**(Cohort B)** | **N = 1227**<br>**(participants in**<br>**Cohort B but not A)** |
| --- | --- | --- | --- | --- |
| **Age (months)** | Median (IQR) | 11.92 (1.08) | 12.00 (1.08) | 12.00 (1.08) |
|  | Range | [10.67, 13.83] | [10.58, 13.67] | [10.58, 13.42] |
| **Sex** | Female | 756 (53.47%) | 1150 (52.90%) | 624 (50.86%) |
|  | Male | 658 (46.53%) | 1024 (47.10%) | 603 (49.14%) |
| **Race/Ethnicity** | White Non-Hispanic | 741 (52.4%) | 1198 (55.11%) | 685 (55.83%) |
|  | Black Non-Hispanic | 190 (13.44%) | 223 (10.26%) | 111 (9.05%) |
|  | Asian Non-Hispanic | 39 (2.76%) | 58 (2.67%) | 27 (2.20%) |
|  | Other Non-Hispanic | 145 (10.25%) | 191 (8.79%) | 103 (8.39%) |
|  | Hispanic | 285 (20.16%) | 477 (21.94%) | 284 (23.15%) |
|  | Missing | 2 (0.14%) | 8 (0.37%) | 6 (0.49%) |
| **BMI** | Median (IQR) | 19.31 (5.36) | 19.06 (5.19) | 19.06 (5.33) |
|  | Missing | 12 (0.85%) | 10 (0.46%) | 4 (0.33%) |
| **Pubertal Stage** | Pre-puberty | 243 (17.19%) | 409 (18.81%) | 247 (20.13%) |
|  | Early puberty | 308 (21.78%) | 453 (20.84%) | 252 (20.54%) |
|  | Mid puberty | 469 (33.17%) | 747 (34.36%) | 419 (34.15%) |
|  | Later puberty | 332 (23.48%) | 470 (21.62%) | 250 (20.37%) |
|  | Missing | 62 (4.38%) | 95 (4.37%) | 59 (4.81%) |
| **Family Income** | <5,000 | 22 (1.56%) | 22 (1.01%) | 17 (1.39%) |
|  | 5,000-24,999 | 68 (4.81%) | 130 (5.98%) | 81 (6.60%) |
|  | 25,000-49,999 | 161 (11.39%) | 233 (10.72%) | 129 (10.51%) |
|  | 50,000-99,999 | 344 (24.33%) | 574 (26.40%) | 342 (27.87%) |
|  | 100,000-199,999 | 482 (34.09%) | 720 (33.12%) | 390 (31.79%) |
|  | >=200,000 | 225 (15.91%) | 347 (15.96%) | 193 (15.73%) |
|  | Missing | 112 (7.92%) | 148 (6.81%) | 75 (6.11%) |
| **Primary Caregiver Education** | Advanced degree (Master’s professional (MD, JD, etc.) and doctoral degrees) | 375 (26.52%) | 580 (26.68%) | 329 (26.81%) |
|  | Bachelor’s degree | 407 (28.78%) | 605 (27.83%) | 338 (27.55%) |
|  | Associate degree | 188 (13.30%) | 299 (13.75%) | 167 (13.61%) |
|  | Some College | 206 (14.57%) | 337 (15.50%) | 201 (16.38%) |
|  | High School/GED | 151 (10.68%) | 209 (9.61%) | 119 (9.70%) |
|  | Did Not Graduate High school | 76 (5.37%) | 126 (5.80%) | 61 (4.97%) |
|  | Missing | 11 (0.78%) | 18 (0.83%) | 12 (0.98%) |

### 2.2 Rapid Response Research (RRR) surveys

Surveys were sent electronically to all eligible youth and caregivers in May, June, August, October, December 2020, and March and May 2021. Youth responses to questions on overall mental health, stress and pandemic-related stress (separate questions) were analyzed following standardization. These questions asked: 1) “How do you think your mental health (emotional well-being) has changed in the past week compared to normal?” [1=much worse to 5=much better]; 2) “COVID-19 presents a lot of uncertainty about the future. In the past 7 days, including today, how stressful have you found this uncertainty to be?” [1=not at all/very slightly to 5=extremely]. An overall stress measure was estimated as a standardized sum of responses from the 4-item Perceived Stress Scale (PSS). Two of these responses–feeling confident in their ability to handle personal problems and feeling things were going their way–were reverse-coded before summing for interpretability. Further, positive affect was calculated as the standardized sum of items from the NIH Toolbox Positive Affect Survey, and negative affect as the standardized sum of items from the NIH Toolbox Emotion Battery (Sadness Survey)^31^. Responses to additional questions on feeling sad, alone, lonely, angry, or scared were also analyzed on a scale from 1=never to 5=almost always. Not all questions were asked across surveys, but four of seven included the question on mental health, and all included the PSS measure of stress^26^. Positive and negative affect were analyzed as t-scores, and all others as z-scores. Response summary statistics are provided in supplemental Table S1.

### 2.3 Mental health variables

Pre-pandemic anxiety, depression, aggressive behaviors, attention problems, internalizing and externalizing behaviors, preference for solitude, and social withdrawal were analyzed. Anxiety, depression, and internalizing and externalizing behaviors were extracted from the Child Behavior Checklist (CBCL) as t-scores. Scores in the range 65-69 are on the clinical borderline and scores ≥70 reflect clinically significant behavioral problems. Preference for solitude and social withdrawal (also extracted from the CBCL) were measured on a Likert scale from 0=not true to 2=very/often true. Prevalence of these problems in the analyzed cohorts and related summary statistics are provided in Table S2.

### 2.4 Other variables

Youth spirituality/religiosity was measured by the response to a question from the Parent Demographics Survey (PDEM) that asked *‘*How important are your child’s religious and spiritual beliefs in his/her daily life?’, on a scale from 1=not at all to 4=very much. Table S16 provides the distribution of responses in the sample of n = 2,641. Parental engagement was measured as the standardized mean of responses to four items from the COVID-19 RRR surveys. These questions assessed parental knowledge of their child’s whereabouts, youth-parent communication, and interaction (parents knowing their child’s plans for the day and eating dinner with them). Responses were coded on a Likert scale from 1=never to 5=always or almost always.

### 2.5 Neuroimaging data analysis

#### 2.5.1 fMRI Preprocessing

Participants in the ABCD study are measured at 21 sites across the US. Neuroimaging data are acquired using 3T Siemens, GE, or Philips scanners. The data are minimally preprocessed before being released^32^. Resting-state fMRI (rs-fMRI) data were analyzed (fMRI acquisition: 2.4 mm isotropic; TR=0.8 s), following processing by the Next Generation Neural Data Analysis platform (NGNDA)^33^, to improve signal-to-noise ratio, suppress artifacts, and harmonize the data across scanners. Voxel-level filtered data (in the frequency range 0.01 to 0.25 Hz) were parcellated and then processed to suppress cardiorespiratory contributions and other artifacts^34^. The highest-quality 5-min run (out of a maximum of 4) was analyzed (which typically had ≤1% of frames censored for motion (median (IQR) = 1.0 (4.0)%).

#### 2.5.2 Estimation of resting-state topological properties

Topological properties of entire brain connectome, individual networks (including large resting-state networks^35^ and the reward^36^, prefrontal cortical, social^37^, and central executive networks^38^), smaller circuits (fronto-thalamic, fronto-amygdala, fronto-basal ganglia, and fronto-striatal circuits), and individual regions (network nodes) were also estimated. These properties included median connectivity (within-and across-network), global (network) and regional clustering, modularity (i.e., community structure), segregation (measured as the ratio of connections within vs outside communities), topological stability (using the largest eigenvalue of the adjacency matrix as a proxy for it), fragility, and robustness, and were estimated using algorithms in NGNDA platform. Analyses involving brain parameters were adjusted for time of day at which participants were scanned, since timing of fMRI data collection may impact network topologies^39^, and percent of fMRI frames censored for motion (relative to scan length).

#### 2.5.3 Structural MRI analysis

Cortical thickness, cortical and subcortical volume, and white matter intensity were estimated from the T1w MRI. These measures are provided by the ABCD at the regional level following brain segmentation using cortical and subcortical atlases, resulting in 98 regions^40^.

### 2.6 Statistical analysis

#### 2.6.1 Mediation model

The model shown in the diagram in supplemental Figure S1 was used to test the hypothesis that pre-pandemic mental health problems directly and indirectly (through their effects on the pre-pandemic organization of brain circuits–the mediator) impacted youth responses during the pandemic. Across paths, linear mixed effect models were developed to account for potential site differences, given substantial differences in state responses and measures to contain the spread of the SARS-CoV-2 virus. Models included a random intercept and slope for site. All parameter p-values were corrected for the False Discovery Rate (FDR)^41^. Mediation analyses were conducted using well-established approaches, and the Sobel test was used to assess the significance of the mediation^42^.

Depending on the path assessed by the model, the dependent or independent variables were either the outcomes from the COVID-19 surveys, individual topological or morphometric brain properties, or pre-pandemic mental health measures. To account for sampling differences between sites, all analyses were adjusted using propensity scores provided by the ABCD. Statistical models also included sex, age, pubertal stage, race-ethnicity, family income, BMI, pandemic onset-to-survey time (months), and scan-to-survey time.

#### 2.6.2 Moderated Multiple Regression (MMR) Models

MMR models^43^ that included interaction terms between individual pre-pandemic mental health issues and individual brain characteristics were also developed to investigate moderation of the relationship between pre-pandemic mental health and pandemic responses by the organization of brain circuits and the morphometric characteristics of their constituent structures. The sign and significance of the interaction term were then examined to assess moderation.

#### 2.6.3 Secondary Analyses

Additional mixed-effects models were used to determine whether protective factors moderated the relationship between pre-pandemic mental health and responses during the pandemic. Parental engagement and youth spirituality were examined as moderators. Finally, models were developed with adjustments for youth responses to COVID survey questions in previous surveys to evaluate the pandemic’s impact over time. Specifically, each model was adjusted in two ways: 1) for the most recent prior survey for an outcome (for example when evaluating associations for stress in survey 3, the response on stress in survey 2 was included in the model), and 2) for the average across all prior surveys for an outcome to capture cumulative effects.

## 3. RESULTS

### 3.1 Associations between Pre-Pandemic Mental Health and Pandemic Responses

About 24.0% of youth had preexisting anxiety, and 7.1% had depression. Pre-pandemic mental health and behavior t-scores were, on average, in the range of 41-51. Only a small fraction of youth had scores ≥65 (∼2-5.5%). Less than 20% preferred solitude at least some times, and ∼6% were socially withdrawn. Summary statistics for pre-pandemic mental health and behavioral issues are summarized in Table S2.

Across both cohorts A and B, several direct associations between existing mental health and behavioral problems and responses to the pandemic stressors were identified. Pre-pandemic anxiety, depression, internalizing, externalizing, and aggressive behaviors were associated with more frequent feelings of anger and loneliness at the May 2020 assessment, i.e., two months following the outbreak (β = 0.09 to 0.18, CI = [0.02, 0.26], p < 0.03). In addition, anxiety, depression, internalizing and/or externalizing behaviors were associated with more frequent sadness, higher stress, and negative affect (β = 0.09 to 0.20, CI = [0.019, 0.30], p < 0.03). Higher pre-pandemic anxiety, depression and internalizing behavior scores were associated with feeling scared in June 2020 (β = 0.13 to 0.21, CI = [0.06, 0.31], p < 0.01), while lower scores and less frequent social withdrawal and preference for solitude were associated with higher positive affect (β = - 0.21 to -0.09, CI = [-0.30, -0.02], p < 0.02). Pre-pandemic mental health issues were also positively associated with feeling alone/lonely, angry, and sad in August 2020 (β = 0.09 to 0.21, CI = [0.01, 0.28], p < 0.05), and aggressive behaviors were also associated with higher stress (β = 0.13 to 0.14 in the two cohorts, CI = [0.02, 0.23], p = 0.04). Anxiety and depression were negatively associated with positive affect (β = -0.21 to -0.11, CI = [-0.31, -0.05], p < 0.01), and positively associated with stress in October 2020 (β = 0.12 to 0.14, CI = [0.03, 0.25], p < 0.04). Finally, in May 2021, i.e., ∼15 months following the outbreak, pre-pandemic aggressive behaviors were positively associated with sadness (β = 0.12 to 0.20 in the two cohorts, CI = [0.06, 0.32], p < 0.01). Detailed model statistics for results that were consistent across cohorts are provided in Table 2. Positive associations between stress in the first ∼7 months of the pandemic (at multiple assessments: May, August, October 2020) and pre-pandemic anxiety and aggressive behaviors are shown in Figure 1a. Negative associations between positive affect during this period (two assessments: June and October 2020) and pre-pandemic anxiety are shown in Figure 1b. Cohort-specific model statistics are summarized in Tables S3 and S4.

**Table 2.**
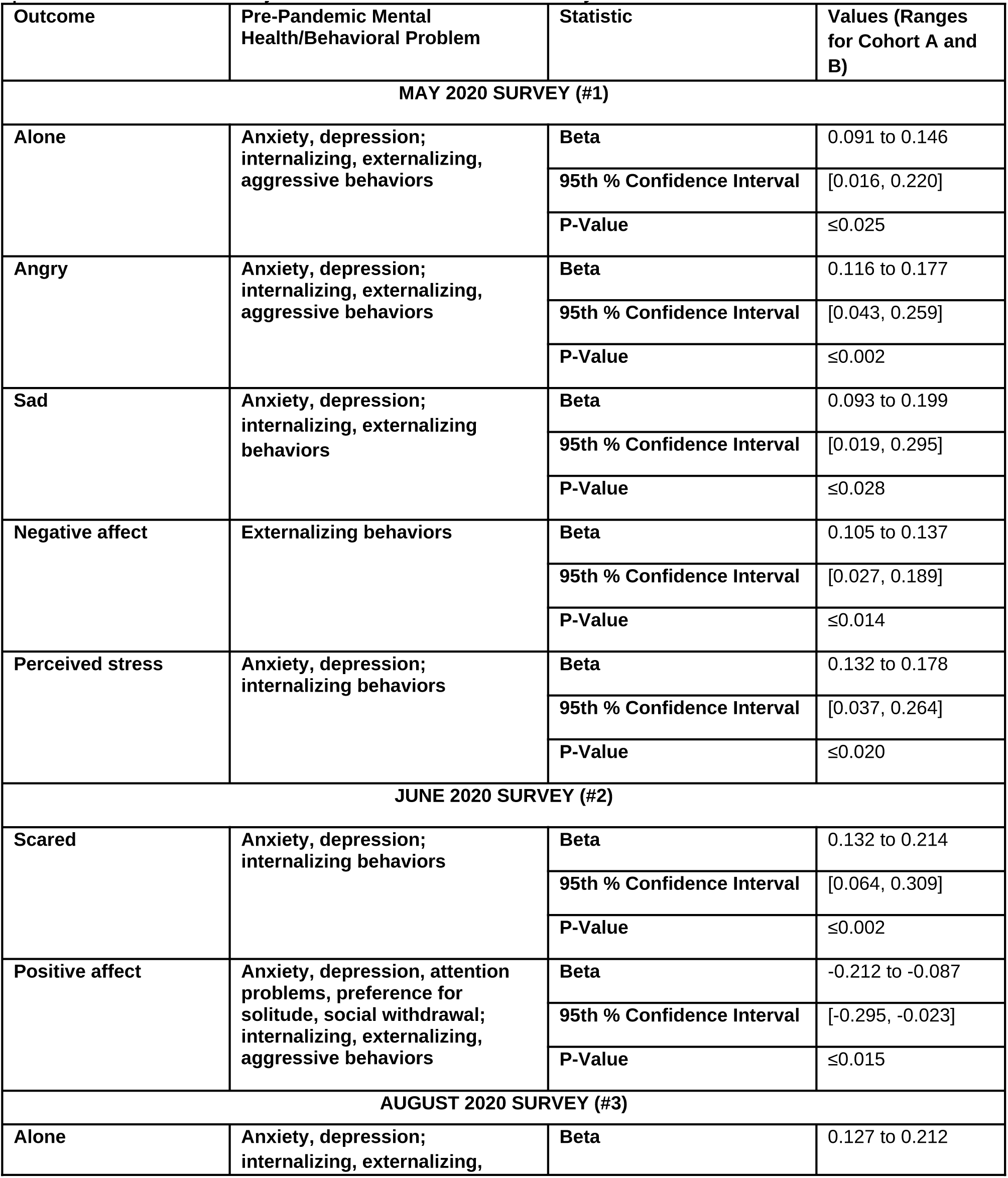
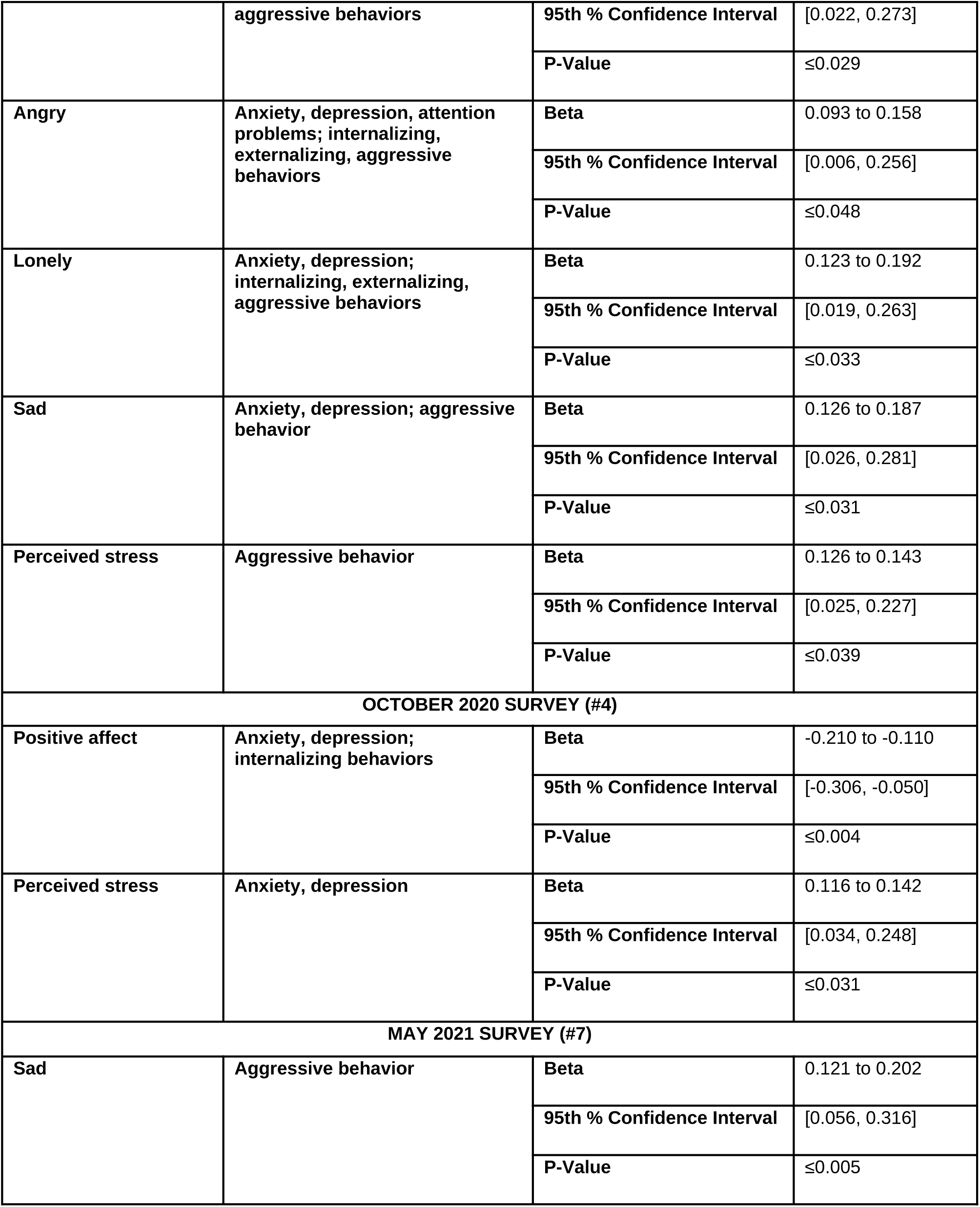
Statistics of models testing predictive relationships between pre-pandemic mental health/behavioral problems and youth survey-based outcomes during the outbreak across both cohorts (Path A of mediation models). Results that are consistent across cohorts A and B are reported. Range of values for both cohorts are provided. All p-values have been adjusted for the False Discovery Rate.

**Figure 1:**
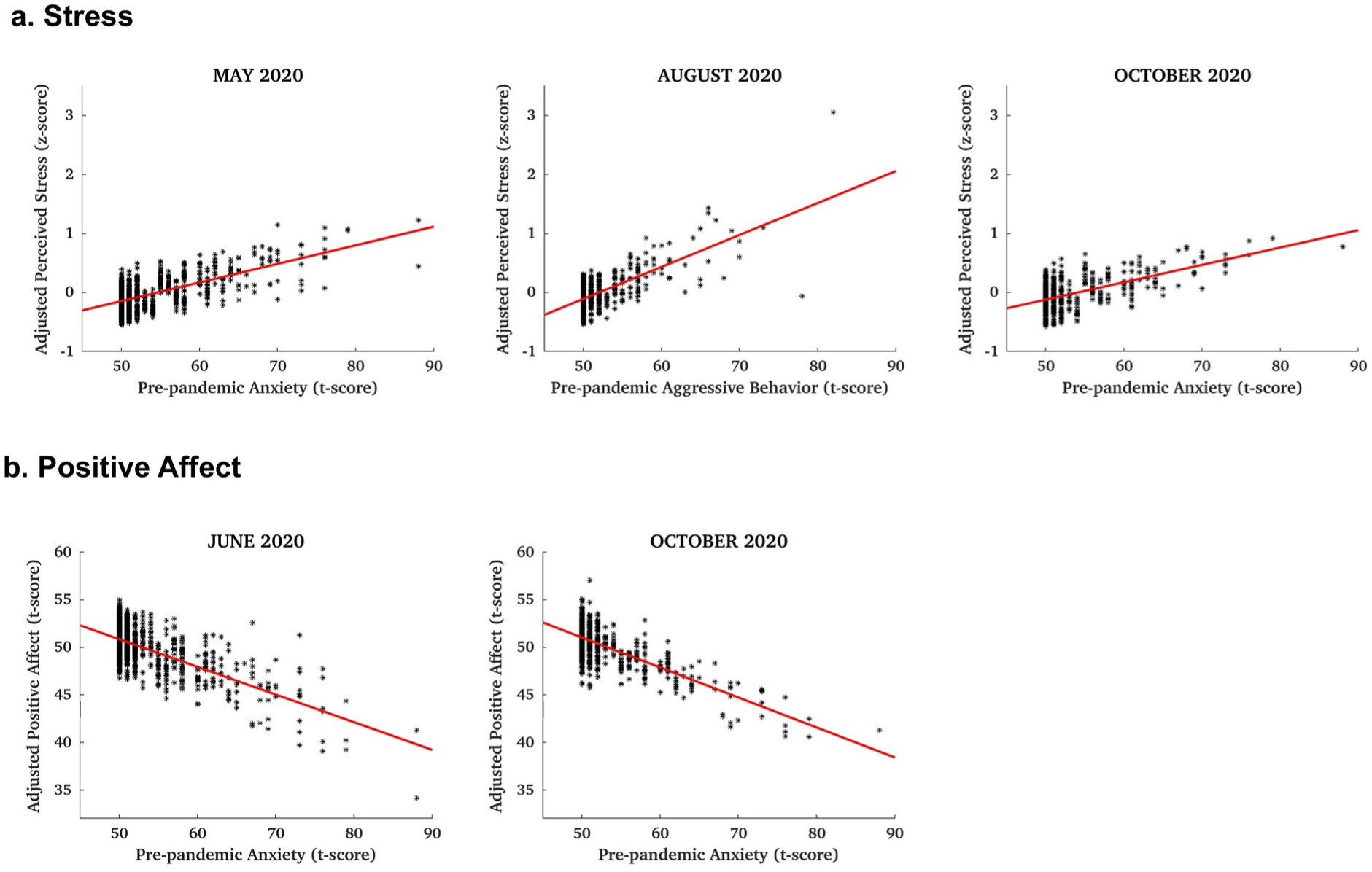
**(a)** Predictive associations between pre-pandemic anxiety (or aggressive behavior) and stress during the first ∼9 months of the pandemic (May, August, October 2020 assessments), and **(b)** corresponding associations with positive affect (June and October 2020 assessments). Stress and positive affect were adjusted for demographic and other participant characteristics, as well as variability across study sites.

Additional associations were estimated between responses at different time points during the first 15 months of the pandemic. Positive affect ∼3 months following the outbreak (June 2020 assessment) was associated with positive affect ∼4 months later (October 2020; β = 0.49 to 0.55, CI = [0.43, 0.65], p < 0.01) in models that were adjusted for pre-pandemic mental health and behavioral issues. Higher negative affect ∼2 months following the outbreak (May 2020 assessment) was associated with higher negative affect ∼3 months later (August 2020, β = 0.68, CI = [0.63, 0.73], p < 0.01), and similarly for feeling alone (β = 0.58, CI = [0.50, 0.61], p < 0.01]). Feeling lonely in the first few months of the pandemic (May and August 2020) was also associated with feeling lonely in December 2020 (β = 0.51, CI = [0.46, 0.57], p < 0.01).

### 3.2 Associations between Pre-Pandemic Mental Health and Brain Characteristics

**Brain circuit characteristics:** In the context of the mediation analysis, associations between pre-pandemic mental health and pre-pandemic brain circuitry were examined in the same samples as those with survey data at each of the seven assessments. In samples from both cohorts A and B, higher anxiety was associated with higher connectivity between the right limbic network and the rest of the brain (β = 0.08 to 0.11, CI = [0.03, 0.19], p < 0.03). Higher attention problems and/or aggressive behavior scores were associated with higher fragility of the left frontoparietal control network (β = 0.08 to 0.13, CI = [0.03, 0.21], p < 0.04), and the right central executive network (β = 0.09 to 0.10, CI = 0.03, 0.17, p < 0.04). Preference for solitude was associated with higher modularity and fragility of the right somatomotor network (β = 0.08 to 0.10, CI = [0.01, 0.18], p < 0.05) and higher segregation of the right temporoparietal network (β = 0.11 to 0.12, CI = [0.04, 0.20], p < 0.05). Model statistics for survey-specific samples are provided in Table 3. Cohort-specific associations between pre-pandemic mental health and topological properties at the connectome and individual network scales are provided in Tables S5-S8. At the regional (node) levels, there were no significant associations in either cohort.

**Table 3.** Statistics of models testing predictive relationships between pre-pandemic mental health/behavioral problems and topological properties of individual networks across both cohorts (Path B of mediation models). Results that are consistent across cohorts A and B are reported. Range of values for both cohorts are provided. All p-values have been adjusted for the False Discovery Rate. CI: Confidence interval.

| MAY 2020 SURVEY (#1) SAMPLE |  |  |  |  |  |
| --- | --- | --- | --- | --- | --- |
| Mental health factor | Network | Properties | Beta | 95th % CI | P-value |
| Anxiety | Limbic (R) | Median connectivity (cross-network) | 0.085 to 0.114 | [0.032, 0.186] | ≤0.023 |
| Attention problems | Frontoparietal control (L) | Fragility | 0.083 to 0.132 | [0.027, 0.205] | ≤0.038 |
| Aggressive behavior | Frontoparietal control (R) | Fragility | 0.083 to 0.130 | [0.028, 0.201] | ≤0.034 |
|  | Central executive (R) | Fragility | 0.085 to 0.096 | [0.027, 0.165] | ≤0.034 |
| Preference for solitude | Somatomotor (R) | Modularity, fragility | 0.079 to 0.098 | [0.009, 0.178] | ≤0.043 |
| JUNE 2020 SURVEY (#2) SAMPLE |  |  |  |  |  |
| Mental health factor | Network | Properties | Beta | 95th % CI | P-value |
| Preference for solitude | Temporoparietal (R) | Segregation | 0.110 to 0.117 | [0.036, 0.198] | ≤0.046 |

**Brain morphometric characteristics:** Higher anxiety was associated with lower thickness of the right temporal gyrus and lower cerebral white matter volume (β = -0.11 to -0.05, CI = [-0.19, -0.001], p < 0.05). Higher depression was associated with higher thickness of the bilateral parahippocampal gyrus and greater white matter intensity in the left temporal pole and right superior frontal gyrus (β = 0.04 to 0.10, CI = [0.01, 0.17], p < 0.05) and lower volume in the right precentral gyrus (β = -0.12 to -0.08, CI = [-0.20, - 0.03], p < 0.04). Attention problems were associated with lower volume of the bilateral rostral anterior cingulate cortex, bilateral cerebellum white matter, right caudal middle frontal gyrus, left pallidum, and left rostral middle frontal and precentral gyri (β = -0.17 to -0.06, CI = [-0.27, -0.002], p < 0.05). Preference for solitude was associated with lower volume of the right nucleus accumbens (β = -0.11 to -0.06, CI = [-0.18, -0.006], p ≤ 0.03) and higher volume of the right hippocampus (β = 0.06 to 0.10, CI = [0.01, 0.18], p < 0.04), while social withdrawal was associated with lower volume of the right paracentral lobule (β = -0.11 to -0.09, CI = [-0.18, -0.03], p < 0.02). Internalizing behaviors were associated with lower volume of the right lateral orbitofrontal cortex and bilateral cerebral white matter (β = -0.14 to -0.05, CI = [-0.23, -0.002], p ≤ 0.04) and higher white matter intensity in the right superior temporal gyrus (β = 0.04 to 0.06, CI = [0.01, 0.10], p ≤ 0.03). Externalizing behaviors were associated with higher thickness of the right pars triangularis and higher white matter intensity in the right temporal pole and right transverse temporal gyrus (β = 0.04 to 0.11, CI = [0.01, 0.17], p < 0.05) and lower volume of bilateral cerebellum and cerebral white matter (β = -0.14 to -0.05, CI = [-0.27, -0.003], p < 0.04). Finally, aggressive behavior was associated with lower white matter volume of the left cerebellum, bilateral cerebral white matter, left hippocampus, and bilateral superior temporal gyrus (β = -0.11 to -0.06, CI = [-0.17, -0.003], p < 0.04). Model statistics for survey-specific samples are provided in Table 4.

**Table 4.**
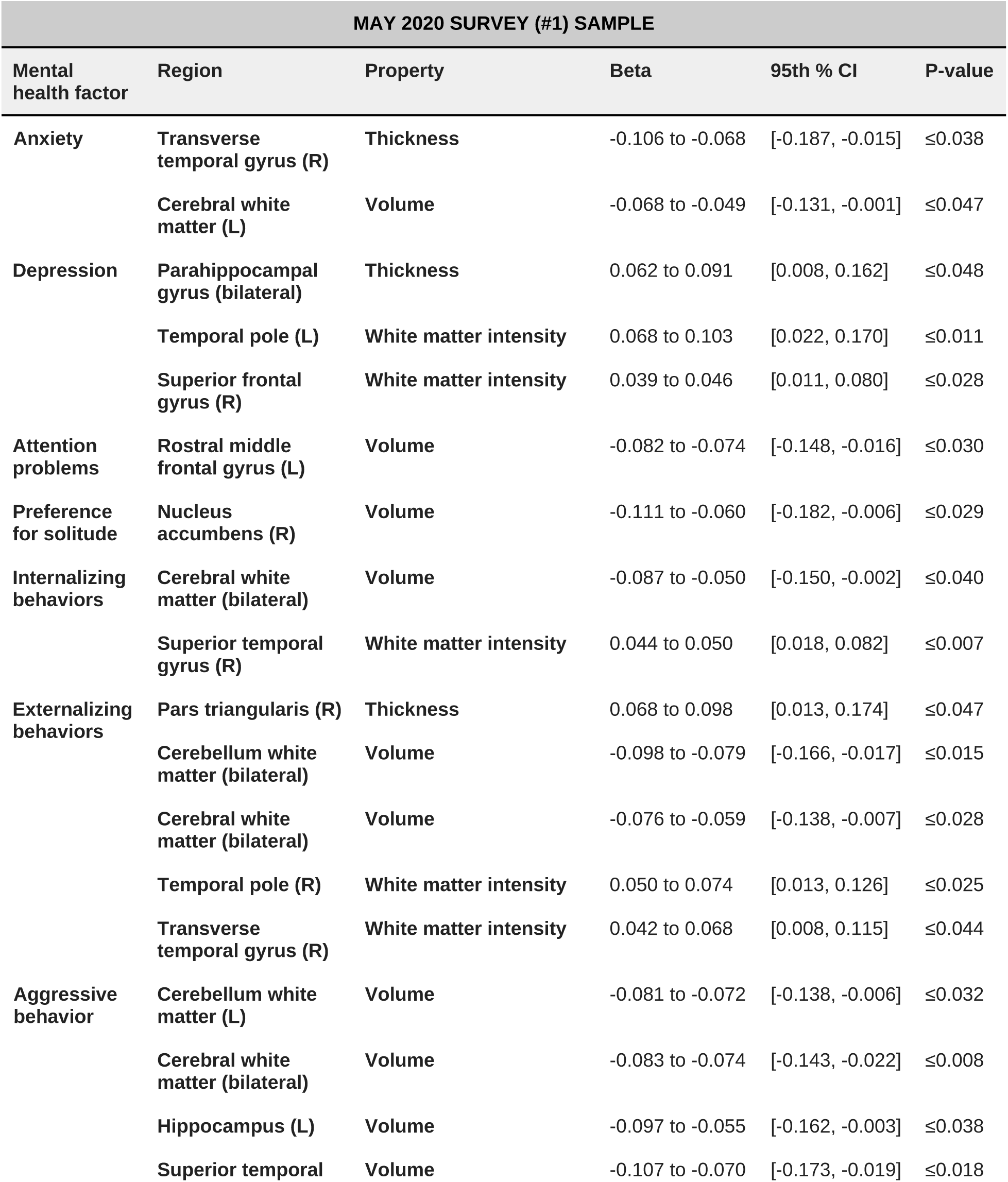
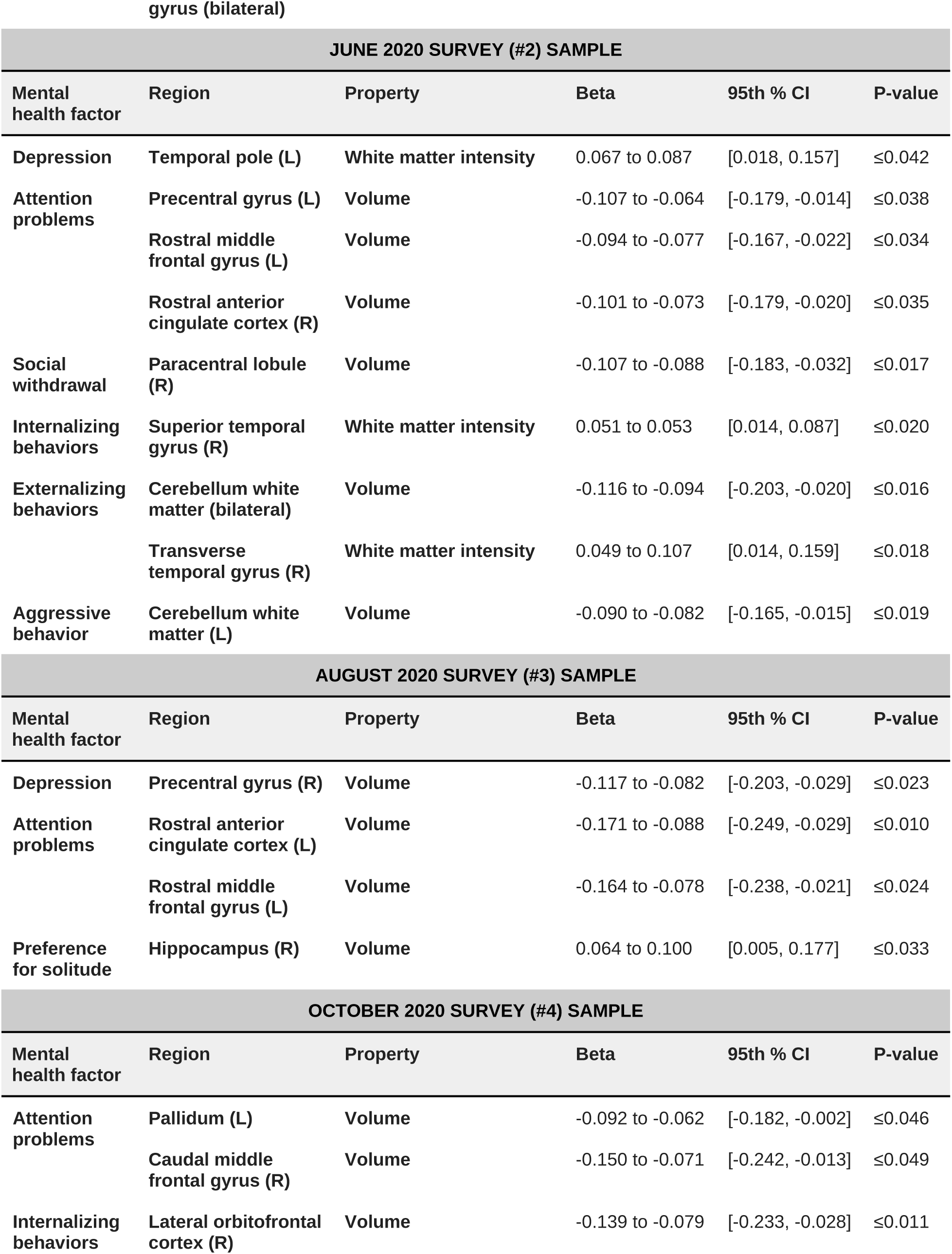
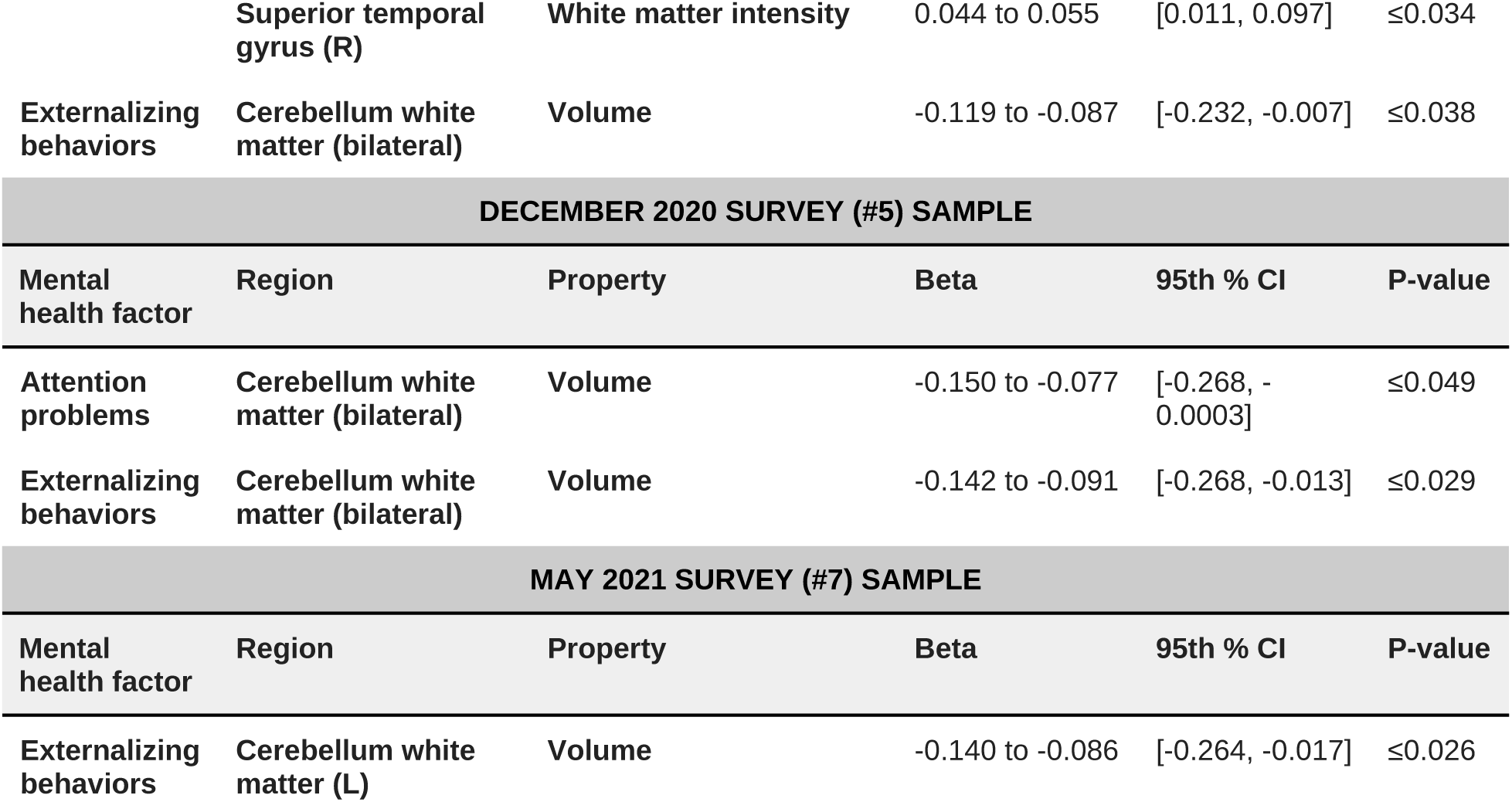
Statistics of models testing predictive relationships between pre-pandemic mental health/behavioral problems and morphometric brain properties across both cohorts (Path B of mediation models). Results that are consistent across cohorts A and B are reported. Range of values for both cohorts are provided. All p-values have been adjusted for the False Discovery Rate. CI: Confidence interval.

| MAY 2020 SURVEY (#1) SAMPLE |  |  |  |  |  |
| --- | --- | --- | --- | --- | --- |
| Mental health factor | Region | Property | Beta | 95th % CI | P-value |
| Anxiety | Transverse temporal gyrus (R) | Thickness | -0.106 to -0.068 | [-0.187, -0.015] | ≤0.038 |
|  | Cerebral white matter (L) | Volume | -0.068 to -0.049 | [-0.131, -0.001] | ≤0.047 |
| Depression | Parahippocampal gyrus (bilateral) | Thickness | 0.062 to 0.091 | [0.008, 0.162] | ≤0.048 |
|  | Temporal pole (L) | White matter intensity | 0.068 to 0.103 | [0.022, 0.170] | ≤0.011 |
|  | Superior frontal gyrus (R) | White matter intensity | 0.039 to 0.046 | [0.011, 0.080] | ≤0.028 |
| Attention problems | Rostral middle frontal gyrus (L) | Volume | -0.082 to -0.074 | [-0.148, -0.016] | ≤0.030 |
| Preference for solitude | Nucleus accumbens (R) | Volume | -0.111 to -0.060 | [-0.182, -0.006] | ≤0.029 |
| Internalizing behaviors | Cerebral white matter (bilateral) | Volume | -0.087 to -0.050 | [-0.150, -0.002] | ≤0.040 |
|  | Superior temporal gyrus (R) | White matter intensity | 0.044 to 0.050 | [0.018, 0.082] | ≤0.007 |
| Externalizing behaviors | Pars triangularis (R) | Thickness | 0.068 to 0.098 | [0.013, 0.174] | ≤0.047 |
|  | Cerebellum white matter (bilateral) | Volume | -0.098 to -0.079 | [-0.166, -0.017] | ≤0.015 |
|  | Cerebral white matter (bilateral) | Volume | -0.076 to -0.059 | [-0.138, -0.007] | ≤0.028 |
|  | Temporal pole (R) | White matter intensity | 0.050 to 0.074 | [0.013, 0.126] | ≤0.025 |
|  | Transverse temporal gyrus (R) | White matter intensity | 0.042 to 0.068 | [0.008, 0.115] | ≤0.044 |
| Aggressive behavior | Cerebellum white matter (L) | Volume | -0.081 to -0.072 | [-0.138, -0.006] | ≤0.032 |
|  | Cerebral white matter (bilateral) | Volume | -0.083 to -0.074 | [-0.143, -0.022] | ≤0.008 |
|  | Hippocampus (L) | Volume | -0.097 to -0.055 | [-0.162, -0.003] | ≤0.038 |
|  | Superior temporal | Volume | -0.107 to -0.070 | [-0.173, -0.019] | ≤0.018 |

gyrus (bilateral)
| JUNE 2020 SURVEY (#2) SAMPLE |  |  |  |  |  |
| --- | --- | --- | --- | --- | --- |
| Mental health factor | Region | Property | Beta | 95th % CI | P-value |
| Depression | Temporal pole (L) | White matter intensity | 0.067 to 0.087 | [0.018, 0.157] | ≤0.042 |
| Attention problems | Precentral gyrus (L) | Volume | -0.107 to -0.064 | [-0.179, -0.014] | ≤0.038 |
|  | Rostral middle frontal gyrus (L) | Volume | -0.094 to -0.077 | [-0.167, -0.022] | ≤0.034 |
|  | Rostral anterior cingulate cortex (R) | Volume | -0.101 to -0.073 | [-0.179, -0.020] | ≤0.035 |
| Social withdrawal | Paracentral lobule (R) | Volume | -0.107 to -0.088 | [-0.183, -0.032] | ≤0.017 |
| Internalizing behaviors | Superior temporal gyrus (R) | White matter intensity | 0.051 to 0.053 | [0.014, 0.087] | ≤0.020 |
| Externalizing behaviors | Cerebellum white matter (bilateral) | Volume | -0.116 to -0.094 | [-0.203, -0.020] | ≤0.016 |
|  | Transverse temporal gyrus (R) | White matter intensity | 0.049 to 0.107 | [0.014, 0.159] | ≤0.018 |
| Aggressive behavior | Cerebellum white matter (L) | Volume | -0.090 to -0.082 | [-0.165, -0.015] | ≤0.019 |
| AUGUST 2020 SURVEY (#3) SAMPLE |  |  |  |  |  |
| Mental health factor | Region | Property | Beta | 95th % CI | P-value |
| Depression | Precentral gyrus (R) | Volume | -0.117 to -0.082 | [-0.203, -0.029] | ≤0.023 |
| Attention problems | Rostral anterior cingulate cortex (L) | Volume | -0.171 to -0.088 | [-0.249, -0.029] | ≤0.010 |
|  | Rostral middle frontal gyrus (L) | Volume | -0.164 to -0.078 | [-0.238, -0.021] | ≤0.024 |
| Preference for solitude | Hippocampus (R) | Volume | 0.064 to 0.100 | [0.005, 0.177] | ≤0.033 |
| OCTOBER 2020 SURVEY (#4) SAMPLE |  |  |  |  |  |
| Mental health factor | Region | Property | Beta | 95th % CI | P-value |
| Attention problems | Pallidum (L) | Volume | -0.092 to -0.062 | [-0.182, -0.002] | ≤0.046 |
|  | Caudal middle frontal gyrus (R) | Volume | -0.150 to -0.071 | [-0.242, -0.013] | ≤0.049 |
| Internalizing behaviors | Lateral orbitofrontal cortex (R) | Volume | -0.139 to -0.079 | [-0.233, -0.028] | ≤0.011 |

|  | Superior temporal gyrus (R) | White matter intensity | 0.044 to 0.055 | [0.011, 0.097] | ≤0.034 |
| --- | --- | --- | --- | --- | --- |
| Externalizing behaviors | Cerebellum white matter (bilateral) | Volume | -0.119 to -0.087 | [-0.232, -0.007] | ≤0.038 |
| DECEMBER 2020 SURVEY (#5) SAMPLE |  |  |  |  |  |
| Mental health factor | Region | Property | Beta | 95th % CI | P-value |
| Attention problems | Cerebellum white matter (bilateral) | Volume | -0.150 to -0.077 | [-0.268, -0.0003] | ≤0.049 |
| Externalizing behaviors | Cerebellum white matter (bilateral) | Volume | -0.142 to -0.091 | [-0.268, -0.013] | ≤0.029 |
| MAY 2021 SURVEY (#7) SAMPLE |  |  |  |  |  |
| Mental health factor | Region | Property | Beta | 95th % CI | P-value |
| Externalizing behaviors | Cerebellum white matter (L) | Volume | -0.140 to -0.086 | [-0.264, -0.017] | ≤0.026 |

### 3.3 Mediation by the Brain

No statistically significant mediation of relationships between pre-pandemic mental health and pandemic responses by the brain’s topological organization were identified. Significant mediation of these relationships by brain morphology was estimated only in individual cohorts and primarily in the first assessment, ∼2 months following the outbreak. In cohort A, the volume of the left insula partially mediated the relationship between pre-pandemic depression and feeling alone in May 2020, the volume of the left superior temporal gyrus partially mediated the relationships between aggressive behaviors and feeling angry, and similarly for externalizing behaviors and anger at the same assessment. White matter volume of the right cerebellum also mediated the relationship between externalizing behaviors and anger in May 2020 (β = 0.10 to 0.14, CI = [0.01, 0.24], p ≤ 0.03). In cohort B, the volume of the right lateral orbitofrontal cortex and left hippocampus partially mediated the relationship between pre-pandemic aggressive behaviors and feeling lonely and angry, respectively, in May 2020 (β = 0.08, CI = [0.01, 0.15], p < 0.04). Model statistics for all mediation results are provided in Tables S9-S10.

### 3.4 Moderation by the Brain

The topological organization of multiple brain networks moderated the relationship between pre-pandemic mental health issues and pandemic responses, primarily in the first ∼3 months following the outbreak. In either cohort A or B, negative associations between pre-pandemic anxiety and depression and overall mental health in May 2020 were attenuated by the strength of connections within the left somatomotor network and between this network and the rest of the brain (β = -0.14 to -0.11, CI = [-0.21, 00.04], p< 0.02). In addition, positive associations between pre-pandemic depression and social withdrawal and stress in May 2020 were attenuated by the community organization (modularity) of the left somatomotor network (β = -0.08, CI = [-0.14, -0.03], p < 0.02). In contrast, strength of connections within the hippocampal network amplified the positive association between pre-pandemic depression and feeling alone in May 2020 (β = 0.11, CI = [0.04, 0.17], p = 0.02). Similar interactions were estimated in June and August 2020. Specifically, modularity of distributed networks, including the prefrontal cortex, frontoparietal control network, and circuits connecting frontal regions to the thalamus, basal ganglia, and/or amygdala attenuated positive relationships between pre-pandemic mental health and behavioral issues and pandemic-related stress in June 2020. In addition, modularity of the right default and central executive networks, and left prefrontal cortex and its connections with the amygdala and the basal ganglia, attenuated positive relationships between pre-pandemic mental health and behavioral problems and feeling scared at the same assessment (β = -0.11 to -0.06, CI = [-0.17, - 0.01], p < 0.05). In contrast, strength of connections within the left temporoparietal network and left hippocampus, amplified associations between pre-pandemic attention problems and pandemic-related stress, and depression and feeling scared, respectively (β = 0.08 to 0.10, CI = [0.02, 0.17], p < 0.03). Finally, modularity of the left somatomotor and left temporoparietal networks attenuated positive associations between pre-pandemic aggressive behaviors, attention problems, and/or social withdrawal and feeling sad and alone in August 2020 (β = -0.12 to -0.07, CI = [-0.17, -0.01], p < 0.04). Detailed model statistics are provided in Table S11.

Relationships between pre-pandemic mental health and behavioral issues and pandemic responses, primarily during the first ∼5-6 months of the pandemic, were also moderated by structural brain characteristics. Higher volume of the right nucleus accumbens, right frontal pole, superior parietal lobule and the hippocampus attenuated relationships between pandemic stress, feeling angry, or feeling alone and pre-pandemic depression, anxiety, and/or internalizing behaviors (β = -0.13 to -0.05, CI = [- 0.24, -0.01], p < 0.05). Also, lower thickness of the right anterior cingulate cortex attenuated the relationship between pre-pandemic internalizing behaviors and pandemic-related stress (β = -0.17, CI = [-0.27, -0.07], p = 0.04). Model statistics for cohorts A and B are summarized in Tables S12 and S13, respectively.

### 3.5 Additional associations

Higher parental engagement was associated with higher positive affect (β = 0.08 to 0.40, CI = [0.03, 0.50], p < 0.01) and lower negative affect, perceived stress, and feelings of sadness, loneliness, anger, and fear (β = -0.32 to -0.10, CI = [-0.40, -0.05], p < 0.01). Parental engagement also moderated relationships between pre-pandemic mental health issues and negative emotions and stress in the first few months following the outbreak, although moderation was not consistent across cohorts. Specifically, it attenuated positive associations between pre-pandemic anxiety and feeling alone, and pre-pandemic aggressive behaviors and feeling angry, as well as negative affect in the first ∼2 months of the pandemic (β = -0.11 to -0.10, CI = [-0.18, -0.03], p < 0.05). Model statistics are provided in Tables S14-S15. In addition, for over 50% of youth, religious beliefs were at least somewhat important in their daily lives (Table S16). Across cohorts, religiosity/spirituality was associated with lower negative affect, less frequent feelings of anger, loneliness, and sadness, and with lower stress, primarily in the first ∼two months of the pandemic, and to a lesser extent ∼5-7 months following the outbreak (β = -0.20 to -0.06, CI = [-0.28, -0.01], p < 0.03). It was also associated with higher positive affect at the October 2020 assessment (β = 0.11 to 0.12 CI = [0.01, 0.22], p < 0.03). Model statistics are summarized in Table S17. Youth from higher-income families reported lower positive affect (β = -0.157 to -0.090, CI = [-0.28, -0.01], p < 0.03) and higher negative affect, including higher sadness, loneliness, anger, and fear (β = 0.08 to 0.22, CI = [0.01, 0.37], p < 0.049). Girls were more likely to report lower positive affect (β = - 2.735 to -0.190, CI = [-4.415, -0.035], p < 0.016) and higher negative affect, as well as higher perceived and pandemic-related stress and higher sadness, loneliness, anger, and fear (β = 0.075 to 4.724, CI = [0.006, 6.117], p < 0.036).

## 4. DISCUSSION

In over 2,600 adolescents, we have investigated direct and indirect impacts of preexisting (pre-pandemic) mental health and behavioral problems on their stress, emotions, and overall mental health during the first ∼15 months of the COVID-19 outbreak. At multiple assessments during this period, especially in the first ∼7 months of the outbreak, pre-pandemic mental health and behavior problems predicted negative emotions and stress, and (to a lesser extent) lower positive affect. Across most assessments, girls reported higher stress, negative affect, and negative emotions, and lower positive affect, in agreement with prior reports of worse mental health and higher stress in girls during the COVID-19 outbreak^20^. Our findings are also in agreement with studies showing that preexisting mental health and behavioral problems were risk factors that amplified the adverse effects of the pandemic on adolescent mental and emotional health^44^.

Youth with preexisting mental health and behavioral problems also had topological and structural brain alterations prior to the pandemic. Specifically, these problems were associated with stronger connections between the limbic network and the rest of the brain, and more topologically fragile frontoparietal control, somatomotor, and central executive networks. Prior studies have linked aberrant connection strength within and between these networks and their topological patterns with mental health problems, including depression and anxiety^45-46^. Topological fragility of these networks may increase risk of miswiring and, in the context of the pandemic, vulnerability to associated stressors. Prior work in the ABCD cohort has shown that lower pre-pandemic topological robustness (higher fragility) of some of these networks were predictive of negative emotions and higher stress during the pandemic^26^. Preexisting mental health problems were also associated with morphometric alterations, especially lower volume of cortical and subcortical regions, including the anterior cingulate cortex, frontal gyrus, orbitofrontal cortex, nucleus accumbens, cerebellum, and overall cerebral white matter. Prior studies have reported associations between lower volume of the anterior cingulate (a brain hub that is widely involved in cognitive function) and mental health problems in children and adolescents^47^, and lower volume of the frontal gyrus (also widely involved in cognitive function, including cognitive control and emotion regulation) and other frontal regions with a similar range of problems, including depression, anxiety, and aggressive behaviors^48-51^. Furthermore, alterations in white matter volume of the cerebellum–which has extensive connections with cortical regions that undergo profound rewiring in adolescence and plays an important role in cognitive (including high-level) function–have been associated with widespread cognitive and mental health issues^52-53^. Of note is that a number of structural alterations in youth with preexisting mental health issues were identified in developing regions that support social function. Social isolation during the pandemic had a profound impact on youth social function^54^, which was likely amplified in youth with an already vulnerable social brain. Together, these findings suggest extensive links between pre-pandemic mental health issues and their topological and morphometric neural substrates, especially in regions supporting cognitive control, emotional regulation and social function, which together increased youth vulnerability to pandemic stressors, leading to higher stress and negative emotions and poor mental health.

To gain a better understanding of the interactions between the brain’s pre-pandemic structural and circuit characteristics, preexisting mental health/behavioral issues, and youth responses during the pandemic, we also examined the brain as a potential mediator of relationships between preexisting mental health and youth pandemic responses. We hypothesized that these preexisting issues impacted youth responses during the pandemic also indirectly, through their neuromodulatory effects. Only morphometric brain characteristics mediated relationships, and only in the early months of the pandemic. Specifically, the volume of the insula, superior temporal gyrus, cerebellum, and orbitofrontal cortex partially mediated associations between pre-pandemic aggressive and/or externalizing behaviors and feelings of anger, and similarly for pre-pandemic depression and feeling lonely in first two months following the outbreak. Together, these structures play important roles in emotional processing and regulation but also social function^37,55-57^. These findings provide mechanistic insights into the brain’s role in how youth responded to the pandemic, especially regions that are central to social function and emotional health, and were already impacted by preexisting mental health issues.

We also examined factors that may positively or negatively moderate the relationships between preexisting mental health issues and stress, emotional and mental health during the pandemic. First, we examined brain characteristics as potential moderators. The organization of multiple networks (including the prefrontal cortex and its subcortical connections, the frontoparietal control, and the somatomotor network) especially their community structure and connection strength attenuated negative associations between preexisting mental health and stress, being scared, sad, and/or lonely during the first ∼2-5 months of the pandemic. Community structure (modularity) is a fundamental topological property of the adult connectome, is associated with its resilience, and increases as a function of neural maturation^58^. Thus, more modular and/or strongly connected brain (and thus more resilient) networks prior to the pandemic may have mitigated the negative impacts of preexisting mental health issues that increased vulnerability to the pandemic stressors. These findings are in agreement with prior work showing that more resilient circuits measured prior to the pandemic were protective for mental and emotional health and stress during the first ∼15 months of the outbreak^26^. In contrast, stronger connections of the left temporoparietal network and the hippocampus were risk factors for emotional health and stress during the pandemic, as they amplified relationships between preexisting behavioral issues and stress and negative emotions (especially feeling scared) in the first few months of the pandemic. Prior research has identified positive associations between connectivity in the hippocampus and stress^59^, and positive associations between connectivity of the temporoparietal network and emotional responses including fear^60^. In addition, a recent study in adolescents and young adults found that stronger connections between the hippocampus and frontal regions, measured prior to the pandemic, were associated with higher stress during the pandemic^61^. Our findings are thus aligned with prior work, and further highlight the critical role of the brain’s organization prior to the pandemic as a protective or risk factor for youth responses to the pandemic’s stressors. Finally, parental engagement and the importance of religious beliefs in the youth’s life were also examined as potential protective factors during the pandemic. Although both were associated with lower stress and negative emotions in the first ∼6-7 months of the pandemic, only parental engagement moderated negative associations between pre-pandemic mental mental health and behavioral issues and negative emotions during the pandemic, and only in the first few months following the outbreak. These findings are aligned with prior studies that have shown that parent involvement and/or religious affiliation were protective factors for adolescent outcomes during the pandemic^25,29^, especially in the first few months of the pandemic, during which social isolation, uncertainty and profound changes in everyday life led to heightened stress and negative emotions.

## Strengths and Limitations

The study has a number of strength, including a large sample size of over 2,600 adolescents, longitudinal surveys assessing youth mental health, stress and emotions during the pandemic, and comprehensive investigation of topological and morphometric brain characteristics, and their complex associations with preexisting mental health and behavior issues, and outcomes during the pandemic. In contrast with previous studies that have focused on direct effects of preexisting mental health issues and youth outcomes during the pandemic, this investigation has allowed us to examine both direct predictive relationships between such preexisting issues and youth responses during the pandemic, but also indirect impacts of these issues through their neuromodulatory effects on underlying neural circuits and structures. It has also allowed us to examine the role of the brain in these predictive relationships, and identify circuits, structures and their characteristics that mediated these relationships and others that moderated the negative effects of preexisting mental health issues on youth pandemic responses. Thus, this investigation has provided mechanistic insights into underlying factors driving (at least partly) the vulnerability of youth with preexisting mental health issues during the pandemic. Despite these strengths, the study also had some limitations. As a retrospective investigation, this study relied on the data provided by the ABCD, and was limited to measures selected by its investigators. In addition, there are potentially many other risk and protective factors that were not measured or investigated, and may have interacted with preexisting mental health problems. The analyses were adjusted for sociodemographic and other individual factors, but there were likely unmeasured, pandemic-related environmental factors that may have affected youth outcomes during the outbreak, and could not be accounted for. Despite these limitations, this study makes a significant contribution towards our incomplete understanding of neural, mental health, and environmental/experiential factors that were either protective for mental health during the COVID-19 pandemic

## 5. CONCLUSIONS

Preexisting anxiety, depression, and/or behavioral problems increased adolescent vulnerability to the COVID-19 pandemic’s stressors, leading to worse mental health, higher stress and negative emotions, directly and through their adverse effects on emotion- and stress-regulatory brain circuits and their constituent structures, especially in girls. Although brain structure characteristics, adversely modulated by preexisting mental health problems, were additional risk factors for negative outcomes, the organization of some brain circuits was protective and attenuated the effects of preexisting mental health issues on youth responses to the pandemic’s stressors. The study identifies adolescent populations who were especially vulnerable during the pandemic (both in terms of their mental health and brain characteristics), who should be followed longitudinally to assess their longer-term (post-pandemic) mental health outcomes. It also identifies specific neural circuits and structures that could be targeted by interventions to improve these outcomes.

## Data Availability

All data used in the present study are publicly available.

https://nda.nih.gov/.

## Abbreviations

MRI: Magnetic Resonance Imaging
fMRI: Functional Magnetic Resonance Imaging

## Acknowledgments

This study was supported by the National Science Foundation, grant #2116707.

