## Supplemental materials for "Pre-pandemic mental health and brain characteristics predict adolescent stress and emotions during the COVID-19 pandemic"

#### 1. SUPPLEMENTAL FIGURES

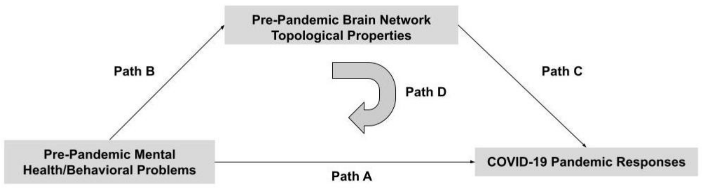


**Figure 1.** Diagram of mediation model used to test the hypothesis that pre-pandemic mental health problems directly and indirectly (through their effects on the pre-pandemic organization of brain circuits - the mediator) impacted youth responses during the pandemic.

### 2. SUPPLEMENTAL TABLES

**Table S1:** COVID Rapid Response Research survey responses, and their distributions in the entire cohort (n=2641).

<table>
<tr><th>Survey response</th><th>Question/Scale Extracted from</th><th>Scale</th><th colspan="3">Median (IQR) of standardized score</th></tr>
<tr><td rowspan="4">Overall mental health<br><br>(z-score)</td><td rowspan="4">“How do you think your mental health (emotional well-being) has improved in the past week compared to normal?”</td><td rowspan="4">1 = much worse;<br>2 = a little worse;<br>3 = about the same;<br>4 = a little better;<br>5 = much better</td><td rowspan="4">Survey</td><td>1</td><td>0.163 (1.224)</td></tr>
<tr><td>3</td><td>0.065 (0.000)</td></tr>
<tr><td>5</td><td>0.186 (1.324)</td></tr>
<tr><td>7</td><td>0.096 (0.000)</td></tr>
<tr><td rowspan="5">Pandemic-uncertainty related Stress (z-score)</td><td rowspan="5">“COVID-19 presents a lot of uncertainty about the future. In the past 7 days, including today, how stressful have you found this uncertainty to be?”</td><td rowspan="5">1 = very slightly or not at all;<br>2 = slightly;<br>3 = moderately;<br>4 = quite a bit;<br>5 = extremely</td><td rowspan="5">Survey</td><td>2</td><td>-0.103 (1.793)</td></tr>
<tr><td>4</td><td>-0.157 (1.783)</td></tr>
<tr><td>5</td><td>-0.134 (1.779)</td></tr>
<tr><td>6</td><td>-0.008 (1.863)</td></tr>
<tr><td>7</td><td>-0.786 (1.022)</td></tr>
<tr><td rowspan="7">Perceived stress<br><br>(z-score)</td><td rowspan="7">4-item Perceived Stress Scale</td><td rowspan="7">Higher score = higher stress level</td><td rowspan="7">Survey</td><td>1</td><td>-0.176 (1.369)</td></tr>
<tr><td>2</td><td>-0.086 (1.425)</td></tr>
<tr><td>3</td><td>-0.0002 (1.430)</td></tr>
<tr><td>4</td><td>-0.187 (1.684)</td></tr>
<tr><td>5</td><td>0.107 (1.357)</td></tr>
<tr><td>6</td><td>-0.207 (1.649)</td></tr>
<tr><td>7</td><td>-0.152 (1.657)</td></tr>
<tr><td rowspan="4">Negative affect<br><br>(t-score)</td><td rowspan="4">NIH Toolbox Emotion Battery (Sadness Survey)</td><td rowspan="4">Higher score = higher occurrence of negative mood</td><td rowspan="4">Survey</td><td>1</td><td>47.246 (13.650)</td></tr>
<tr><td>3</td><td>46.510 (15.488)</td></tr>
<tr><td>5</td><td>47.813 (15.037)</td></tr>
<tr><td>7</td><td>47.999 (14.918)</td></tr>
</table>

| | | | | | |
|---|---|---|---|---|---|
| **Positive affect**<br><br>**(t-score)** | NIH Toolbox Positive Affect Survey | Higher score = higher occurrence of positive mood | **Survey** | **2** | 50.87 (14.927) |
| | | | | **4** | 49.363 (16.305) |
| | | | | **6** | 48.831 (16.256) |
| **Felt sad**<br><br>**(z-score)** | "In the past week, I felt sad" | 1 = never;<br>2 = almost never;<br>3 = sometimes;<br>4 = often;<br>5 = almost always | **Survey** | **1** | -0.085 (1.910) |
| | | | | **3** | -0.002 (1.989) |
| | | | | **5** | -0.176 (1.823) |
| | | | | **7** | -0.153 (1.818) |
| **Felt alone**<br><br>**(z-score)** | "In the past week, I felt alone" | 1 = never;<br>2 = almost never;<br>3 = sometimes;<br>4 = often;<br>5 = almost always | **Survey** | **1** | -0.778 (0.947) |
| | | | | **3** | -0.752 (1.004) |
| | | | | **5** | 0.081 (1.816) |
| | | | | **7** | 0.091 (1.825) |
| **Felt lonely**<br><br>**(z-score)** | "In the past week, I felt lonely" | 1 = never;<br>2 = almost never;<br>3 = sometimes;<br>4 = often;<br>5 = almost always | **Survey** | **1** | -0.015 (1.809) |
| | | | | **3** | 0.100 (1.909) |
| | | | | **5** | -0.055 (1.736) |
| | | | | **7** | -0.023 (1.776) |
| **Felt angry**<br><br>**(z-score)** | "In the past week, I felt angry or frustrated" | 1 = never;<br>2 = almost never;<br>3 = sometimes;<br>4 = often;<br>5 = almost always | **Survey** | **1** | -0.143 (1.861) |
| | | | | **3** | -0.044 (1.991) |
| | | | | **5** | -0.216 (1.817) |
| | | | | **7** | -0.210 (1.801) |
| **Felt scared**<br><br>**(z-score)** | "In the past week, I felt scared" | 1 = never;<br>2 = almost never;<br>3 = sometimes;<br>4 = often;<br>5 = almost always | **Survey** | **2** | 0.210 (1.165) |

**Table S2.** Prevalence and summary statistics (median and interquartile range (IQR) of pre-pandemic depression, anxiety, and behavioral problems in cohorts A and B.

| | Statistic | Cohort A (n = 1414) | Cohort B (n = 2174) |
|---|---|---|---|
| **Depression** | Median (IQR) | 50 (5) | 50 (4) |
| | Prevalence [N (%)] (scores ≥65) | 78 (5.52%) | 97 (4.46%) |
| | Prevalence [N (%)] (scores ≥70) | 36 (2.55%) | 39 (1.79%) |
| **Anxiety** | Median (IQR) | 51 (3) | 51 (2) |
| | Prevalence [N (%)] (scores ≥65) | 69 (4.88%) | 98 (4.51%) |
| | Prevalence [N (%)] (scores ≥70) | 27 (1.91%) | 40 (1.84%) |
| **Aggressive behaviors** | Median (IQR) | 50 (1) | 50 (1) |
| | Prevalence [N (%)] (scores ≥65) | 31 (2.19%) | 39 (1.79%) |
| | Prevalence [N (%)] (scores ≥70) | 13 (0.92%) | 13 (0.60%) |
| **Attention problems** | Median (IQR) | 51 (3) | 50 (3) |
| | Prevalence [N(%)] (scores ≥65) | 39 (2.76%) | 45 (2.07%) |
| | Prevalence (%) [N (%)] (scores ≥70) | 17 (1.20%) | 15 (0.69%) |
| **Internalizing behaviors** | Median (IQR) | 47 (15) | 46 (14) |
| | Prevalence (scores ≥65) | 61 (4.31%) | 107 (4.92%) |
| **Externalizing behaviors** | Median (IQR) | 41 (15) | 41 (15) |
| | Prevalence [N (%)] (scores ≥65) | 26 (1.84%) | 28 (1.29%) |
| **Preference for solitude** | Not True | 1134 (80.20%) | 1781 (81.92%) |
| | Somewhat OR Very True | 279 (19.73%) | 391 (17.99%) |
| **Social withdrawal** | Not True | 1327 (93.85%) | 2047 (94.16%) |
| | Somewhat OR Very True | 86 (6.09%) | 125 (5.75%) |

**Table S3.** Statistics of models testing predictive relationships between pre-pandemic mental health/behavioral problems and youth survey-based outcomes during the first ~15 months of the pandemic in Cohort A (Path A of mediation models). All p-values have been adjusted for the False Discovery Rate.

| Outcome | Pre-Pandemic Mental Health/Behavioral Problem | Statistic | Value |
|---|---|---|---|
| **MAY 2020 SURVEY (#1)** | | | |
| **Alone** | **Anxiety, depression, social withdrawal; internalizing, externalizing, aggressive behaviors** | **Beta** | 0.077 to 0.133 |
| | | **95th % Confidence Interval** | [0.006, 0.220] |
| | | **P-Value** | ≤0.039 |
| **Angry** | **Anxiety, depression; internalizing, externalizing, aggressive behaviors** | **Beta** | 0.128 to 0.177 |
| | | **95th % Confidence Interval** | [0.049, 0.259] |
| | | **P-Value** | ≤0.002 |
| **Sad** | **Anxiety, depression; internalizing, externalizing behaviors** | **Beta** | 0.093 to 0.199 |
| | | **95th % Confidence Interval** | [0.019, 0.295] |
| | | **P-Value** | ≤0.028 |
| **Negative affect** | **Anxiety, depression; internalizing, externalizing, aggressive behaviors** | **Beta** | 0.093 to 0.192 |
| | | **95th % Confidence Interval** | [0.023, 0.280] |
| | | **P-Value** | ≤0.016 |
| **Perceived stress** | **Anxiety, depression, social withdrawal; internalizing behaviors** | **Beta** | 0.089 to 0.178 |
| | | **95th % Confidence Interval** | [0.013, 0.264] |
| | | **P-Value** | ≤0.043 |
| **JUNE 2020 SURVEY (#2)** | | | |
| **Scared** | **Anxiety, depression; internalizing behaviors** | **Beta** | 0.149 to 0.214 |
| | | **95th % Confidence Interval** | [0.064, 0.309] |
| | | **P-Value** | ≤0.002 |
| **Positive affect** | **Anxiety, depression, attention problems, preference for solitude, social withdrawal; internalizing, externalizing, aggressive behaviors** | **Beta** | -0.212 to -0.111 |
| | | **95th % Confidence Interval** | [-0.295, -0.023] |
| | | **P-Value** | ≤0.015 |
| **AUGUST 2020 SURVEY (#3)** | | | |
| **Alone** | **Anxiety, depression; internalizing, externalizing, aggressive behaviors** | **Beta** | 0.127 to 0.186 |
| | | **95th % Confidence Interval** | [0.022, 0.273] |
| | | **P-Value** | ≤0.029 |
| **Angry** | **Anxiety, depression, attention problems; internalizing, externalizing, aggressive behaviors** | **Beta** | 0.093 to 0.151 |
| | | **95th % Confidence Interval** | [0.006, 0.256] |

| | | | |
|---|---|---|---|
| | | **P-Value** | ≤0.048 |
| **Lonely** | **Anxiety, depression; internalizing, externalizing, aggressive behaviors** | **Beta** | 0.123 to 0176 |
| | | **95th % Confidence Interval** | [0.019, 0.263] |
| | | **P-Value** | ≤0.033 |
| **Sad** | **Anxiety, depression; aggressive behavior** | **Beta** | 0.132 to 0.187 |
| | | **95th % Confidence Interval** | [0.026, 0.281] |
| | | **P-Value** | ≤0.031 |
| **Negative affect** | **Anxiety, depression; internalizing, externalizing, aggressive behaviors** | **Beta** | 0.128 to 0.184 |
| | | **95th % Confidence Interval** | [0.024, 0.281] |
| | | **P-Value** | ≤0.026 |
| **Perceived stress** | **Aggressive behavior** | **Beta** | 0.126 |
| | | **95th % Confidence Interval** | [0.025, 0.227] |
| | | **P-Value** | ≤0.039 |
| **OCTOBER 2020 SURVEY (#4)** | | | |
| **Positive affect** | **Anxiety, depression; internalizing behaviors** | **Beta** | -0.210 to -0.168 |
| | | **95th % Confidence Interval** | [-0.306, -0.066] |
| | | **P-Value** | ≤0.004 |
| **Perceived stress** | **Anxiety, depression** | **Beta** | 0.136 to 0.142 |
| | | **95th % Confidence Interval** | [0.034, 0.248] |
| | | **P-Value** | ≤0.031 |
| **Pandemic-related stress** | **Anxiety** | **Beta** | 0.142 |
| | | **95th % Confidence Interval** | [0.043, 0.24] |
| | | **P-Value** | ≤0.039 |
| **DECEMBER 2020 SURVEY (#5)** | | | |
| **Overall mental health** | **Depression** | **Beta** | -0.276 |
| | | **95th % Confidence Interval** | [-0.412, -0.141] |
| | | **P-Value** | <0.001 |
| **MAY 2021 SURVEY (#7)** | | | |
| **Angry** | **Aggressive behavior** | **Beta** | 0.215 |
| | | **95th % Confidence Interval** | [0.102, 0.329] |
| | | **P-Value** | <0.001 |

| | | | |
|---|---|---|---|
| **Sad** | **Aggressive behavior** | **Beta** | 0.202 |
| | | **95th % Confidence Interval** | [0.087, 0.316] |
| | | **P-Value** | ≤0.005 |

**Table S4. Statistics of models testing predictive relationships between pre-pandemic mental health/behavioral problems and youth survey-based outcomes during the first ~15 months of the pandemic in Cohort B (Path A of mediation models). All p-values have been adjusted for the False Discovery Rate.**

| Outcome | Pre-Pandemic Mental Health/Behavioral Problem | Statistic | Value |
|---|---|---|---|
| MAY 2020 SURVEY (#1) | | | |
| Alone | Anxiety, depression; internalizing, externalizing, aggressive behaviors | Beta | 0.095 to 0.146 |
| | | 95th % Confidence Interval | [0.029, 0.198] |
| | | P-Value | ≤0.005 |
| Angry | Anxiety, depression; internalizing, externalizing, aggressive behaviors | Beta | 0.116 to 0.141 |
| | | 95th % Confidence Interval | [0.043, 0.199] |
| | | P-Value | ≤0.002 |
| Lonely | Anxiety, depression; internalizing, externalizing, aggressive behaviors | Beta | 0.079 to 0.136 |
| | | 95th % Confidence Interval | [0.025, 0.188] |
| | | P-Value | ≤0.004 |
| Sad | Anxiety, depression, attention problems, social withdrawal; internalizing, externalizing, aggressive behaviors | Beta | 0.064 to 0.180 |
| | | 95th % Confidence Interval | [0.004, 0.237] |
| | | P-Value | ≤0.037 |
| Negative affect | Anxiety, depression, attention problems; internalizing, externalizing, aggressive behaviors | Beta | 0.069 to 0.180 |
| | | 95th % Confidence Interval | [0.016, 0.233] |
| | | P-Value | ≤0.010 |
| Perceived stress | Anxiety, depression; internalizing, externalizing, aggressive behaviors | Beta | 0.075 to 0.176 |
| | | 95th % Confidence Interval | [0.013, 0.236] |
| | | P-Value | ≤0.018 |
| JUNE 2020 SURVEY (#2) | | | |
| Scared | Anxiety, depression, attention problems, preference for solitude, social withdrawal; internalizing, aggressive behaviors | Beta | 0.056 to 0.173 |
| | | 95th % Confidence Interval | [0.003, 0.226] |
| | | P-Value | ≤0.038 |
| Felt something awful might happen | Depression, anxiety, attention problems; internalizing behaviors | Beta | 0.113 to 0.176 |
| | | 95th % Confidence Interval | [0.058, 0.231] |
| | | P-Value | <0.001 |
| Positive affect (t-score) | Anxiety, depression, attention problems, preference for solitude, social withdrawal; internalizing, externalizing, aggressive behaviors | Beta | -0.191 to -0.087 |
| | | 95th % Confidence Interval | [-0.253, -0.025] |
| | | P-Value | ≤0.006 |
| Perceived stress | Depression, anxiety, attention problems; internalizing, aggressive behaviors | Beta | 0.096 to 0.169 |
| | | 95th % Confidence Interval | [0.044, 0.223] |
| | | P-Value | <0.001 |

| | | | |
|---|---|---|---|
| **Pandemic-related stress** | **Depression, anxiety, attention problems; internalizing behaviors** | **Beta** | 0.080 to 0.114 |
| | | **95th % Confidence Interval** | [0.015, 0.173 |
| | | **P-Value** | ≤0.015 |
| **AUGUST 2020 SURVEY (#3)** | | | |
| **Alone** | **Anxiety, depression, attention problems, preference for solitude, social withdrawal; internalizing, externalizing, aggressive behaviors** | **Beta** | 0.075 to 0.212 |
| | | **95th % Confidence Interval** | [0.018, 0.270] |
| | | **P-Value** | ≤0.010 |
| **Angry** | **Anxiety, depression, attention problems; internalizing, externalizing, aggressive behaviors** | **Beta** | 0.106 to 0.158 |
| | | **95th % Confidence Interval** | [0.032, 0.225] |
| | | **P-Value** | ≤0.005 |
| **Lonely** | **Anxiety, depression, attention problems; internalizing, externalizing, aggressive behaviors** | **Beta** | 0.104 to 0.192 |
| | | **95th % Confidence Interval** | [0.038, 0.251] |
| | | **P-Value** | ≤0.002 |
| **Sad** | **Anxiety, depression, attention problems, preference for solitude, social withdrawal; internalizing, externalizing, aggressive behavior** | **Beta** | 0.060 to 0.197 |
| | | **95th % Confidence Interval** | [0.003, 0.253] |
| | | **P-Value** | ≤0.040 |
| **Negative affect** | **Anxiety, depression, attention problems, preference for solitude, social withdrawal; internalizing, externalizing, aggressive behaviors** | **Beta** | 0.059 to 0.208 |
| | | **95th % Confidence Interval** | [0.003, 0.264] |
| | | **P-Value** | ≤0.038 |
| **Perceived stress** | **Depression, anxiety, attention problems; aggressive behavior** | **Beta** | 0.106 to 0.172 |
| | | **95th % Confidence Interval** | [0.034, 0.238] |
| | | **P-Value** | ≤0.004 |
| **OCTOBER 2020 SURVEY (#4)** | | | |
| **Positive affect** | **Anxiety, depression, attention problems, preference for solitude, social withdrawal; internalizing, externalizing, aggressive behaviors** | **Beta** | -0.179 to -0.110 |
| | | **95th % Confidence Interval** | [-0.238, -0.044] |
| | | **P-Value** | ≤0.002 |
| **Perceived stress** | **Anxiety, depression, attention problems, social withdrawal; internalizing, externalizing, aggressive behaviors** | **Beta** | 0.084 to 0.133 |
| | | **95th % Confidence Interval** | [0.016, 0.195] |
| | | **P-Value** | ≤0.016 |
| **DECEMBER 2020 SURVEY (#5)** | | | |
| **Alone** | **Depression; internalizing, externalizing, aggressive** | **Beta** | 0.077 to 0.116 |

| | | | |
|---|---|---|---|
| | behaviors | 95th % Confidence Interval | [0.014, 0.177] |
| | | P-Value | ≤0.017 |
| Angry | Depression; externalizing, aggressive behaviors | Beta | 0.080 to 0.125 |
| | | 95th % Confidence Interval | [0.013, 0.194] |
| | | P-Value | ≤0.019 |
| Lonely | Depression; internalizing, externalizing, aggressive behaviors | Beta | 0.086 to 0.132 |
| | | 95th % Confidence Interval | [0.022, 0.192] |
| | | P-Value | ≤0.013 |
| Sad | Internalizing, externalizing behaviors | Beta | 0.094 to 0.098 |
| | | 95th % Confidence Interval | [0.034, 0.157] |
| | | P-Value | ≤0.002 |
| Negative affect | Depression; internalizing, externalizing, aggressive behaviors | Beta | 0.088 to 0.120 |
| | | 95th % Confidence Interval | [0.026, 0.179] |
| | | P-Value | ≤0.005 |
| Perceived stress | Depression, anxiety, social withdrawal; internalizing, externalizing, aggressive behaviors | Beta | 0.090 to 0.124 |
| | | 95th % Confidence Interval | [0.024, 0.185] |
| | | P-Value | ≤0.007 |
| MARCH 2021 SURVEY (#6) | | | |
| Positive affect | Depression, anxiety | Beta | -0.121 to -0.108 |
| | | 95th % Confidence Interval | [-0.184, -0.040] |
| | | P-Value | ≤0.002 |
| Perceived stress | Depression, anxiety, attention problems, preference for solitude, social withdrawal; internalizing, externalizing, aggressive behaviors | Beta | 0.070 to 0.149 |
| | | 95th % Confidence Interval | [0.006, 0.206] |
| | | P-Value | ≤0.032 |
| MAY 2021 SURVEY (#7) | | | |
| Lonely | Depression, anxiety, attention problems; internalizing, externalizing, aggressive behaviors | Beta | 0.086 to 0.162 |
| | | 95th % Confidence Interval | [0.007, 0.223] |
| | | P-Value | ≤0.034 |
| Sad | Depression, anxiety, attention problems; aggressive behavior | Beta | 0.111 to 0.133 |
| | | 95th % Confidence Interval | [0.033, 0.196] |
| | | P-Value | ≤0.005 |
| Negative affect | Attention problems; aggressive behavior | Beta | 0.108 to 0.127 |

| | | | |
|---|---|---|---|
| | | **95th % Confidence Interval** | [0.021, 0.200] |
| | | **P-Value** | ≤0.015 |
| **Perceived stress** | **Depression, anxiety, attention problems, preference for solitude, social withdrawal; internalizing, externalizing, aggressive behaviors** | **Beta** | 0.085 to 0.181 |
| | | **95th % Confidence Interval** | [0.024, 0.241] |
| | | **P-Value** | ≤0.009 |

**Table S5.** Statistics of models testing predictive relationships between pre-pandemic mental health/behavioral problems and whole-brain topological properties in Cohort A (Path B of mediation models). All p-values have been adjusted for the False Discovery Rate. CI: Confidence interval.

| MAY 2020 SURVEY (#1) SAMPLE | | | | |
|---|---|---|---|---|
| **Mental health factor** | **Properties** | **Beta** | **95th % CI** | **P-value** |
| **Preference for solitude** | **Global efficiency** | -0.127 | [-0.196, -0.058] | <0.001 |
| | **Global clustering** | -0.086 | [-0.156, -0.016] | 0.016 |
| | **Modularity** | 0.117 | [0.043, 0.190] | 0.002 |
| | **Fragility** | 0.084 | [0.011, 0.158] | 0.025 |
| **AUGUST 2020 SURVEY (#3) SAMPLE** | | | | |
| **Mental health factor** | **Properties** | **Beta** | **95th % CI** | **P-value** |
| **Attention problems** | **Fragility** | 0.152 | [0.064, 0.239] | <0.001 |

**Table S6.** Statistics of models testing predictive relationships between pre-pandemic mental health/behavioral problems and whole-brain topological properties in Cohort B (Path B of mediation models). All p-values have been adjusted for the False Discovery Rate. CI: Confidence interval.

| OCTOBER 2020 SURVEY (#4) SAMPLE | | | | |
|---|---|---|---|---|
| **Mental health factor** | **Properties** | **Beta** | **95th % CI** | **P-value** |
| **Preference for solitude** | **Global efficiency** | -0.084 | [-0.140, -0.028] | 0.032 |
| | **Global clustering** | -0.068 | [-0.129, -0.007] | 0.048 |
| | **Modularity** | 0.071 | [0.009, 0.132] | 0.048 |
| | **Segregation** | 0.070 | [0.013, 0.128] | 0.048 |

**Table S7.** Statistics of models testing predictive relationships between pre-pandemic mental health/behavioral problems and topological properties of individual networks in Cohort A (Path B of mediation models). All p-values have been adjusted for the False Discovery Rate. CI: Confidence interval.

| MAY 2020 SURVEY (#1) SAMPLE | | | | | |
|---|---|---|---|---|---|
| **Mental health factor** | **Network** | **Properties** | **Beta** | **95th % CI** | **P-value** |
| **Anxiety** | **Limbic (R)** | **Median connectivity (cross-network)** | 0.114 | [0.041, 0.186] | 0.023 |
| | **Thalamus** | **Median connectivity (cross-network)** | 0.108 | [0.037, 0.179] | 0.029 |
| **Attention problems** | **Frontoparietal control (L)** | **Fragility** | 0.132 | [0.060, 0.205] | 0.004 |
| **Aggressive behavior** | **Dorsal attention (L)** | **Topological robustness, global efficiency, topological stability** | -0.086 to -0.083 | [-0.155, -0.014] | ≤0.047 |
| | **Frontoparietal control (bilateral)** | **Topological robustness, global efficiency, topological stability** | -0.099 to -0.081 | [-0.167, -0.014] | ≤0.035 |
| | | **Fragility** | 0.111 to 0.130 | [0.041, 0.201] | ≤0.010 |
| | **Central executive (R)** | **Topological robustness, topological stability, global efficiency, global clustering** | -0.081 to -0.076 | [-0.148, -0.008] | ≤0.048 |
| | | **Fragility** | 0.096 | [0.027, 0.165] | 0.034 |
| | **Cerebellum (L)** | **Fragility** | 0.101 | [0.032, 0.171] | 0.044 |
| | **Fronto-striatal-thalamic circuit (R)** | **Fragility** | 0.107 | [0.037, 0.177] | 0.030 |
| **Preference for solitude** | **Dorsal attention (bilateral)** | **Topological robustness, topological stability, global efficiency, global clustering** | -0.135 to -0.079 | [-0.205, -0.007] | ≤0.049 |
| | | **Modularity, fragility** | 0.087 to 0.117 | [0.013, 0.193] | ≤0.044 |
| | **Dorsal attention (R)** | **Segregation** | 0.095 | [0.026, 0.165] | 0.013 |
| | **Control (bilateral)** | **Topological robustness, topological stability, global efficiency** | -0.120 to -0.081 | [-0.188, -0.012] | ≤0.043 |
| | | **Modularity** | 0.083 to 0.160 | [0.013, 0.231] | ≤0.043 |
| | **Control (L)** | **Global clustering** | -0.088 | [-0.158, -0.017] | 0.025 |
| | | **Fragility** | 0.160 | [0.089, 0.231] | <0.001 |
| | **Frontoparietal control (R)** | **Segregation** | 0.097 | [0.026, 0.169] | 0.043 |
| | **Temporoparietal (L)** | **Topological robustness, topological stability, global efficiency, global clustering** | -0.120 to -0.097 | [-0.193, -0.023] | ≤0.020 |

| | | | | |
|---|---|---|---|---|
| | **Modularity** | 0.099 | [0.024, 0.173] | 0.020 |
| **Temporoparietal (R)** | **Segregation** | 0.127 | [0.054, 0.200] | 0.007 |
| **Reward (bilateral)** | **Global efficiency** | -0.103 to -0.101 | [-0.169, -0.033] | ≤0.035 |
| **Reward (R)** | **Topological robustness, topological stability, global clustering** | -0.088 to -0.078 | [-0.167, -0.010] | ≤0.046 |
| | **Modularity, segregation** | 0.084 | [0.010, 0.158] | 0.046 |
| **Social (bilateral)** | **Global efficiency** | -0.103 to -0.079 | [-0.171, -0.011] | ≤0.038 |
| | **Segregation** | 0.104 | [0.032, 0.175] | 0.037 |
| **Social (R)** | **Topological robustness, topological stability, global clustering** | -0.087 to -0.079 | [-0.155, -0.008] | ≤0.041 |
| | **Modularity, fragility** | 0.082 to 0.093 | [0.011, 0.163] | ≤0.038 |
| **Prefrontal cortex (bilateral)** | **Topological robustness, topological stability, global efficiency** | -0.117 to -0.087 | [-0.184, -0.018] | ≤0.034 |
| **Prefrontal cortex (L)** | **Global clustering** | -0.091 | [-0.158, -0.024] | 0.033 |
| **Prefrontal cortex (R)** | **Modularity, fragility** | 0.107 to 0.111 | [0.043, 0.180] | ≤0.013 |
| **Fronto-thalamic circuit (bilateral)** | **Global efficiency** | -0.110 to -0.106 | [-0.176, -0.041] | ≤0.015 |
| **Fronto-thalamic circuit (R)** | **Topological robustness, global clustering, topological stability** | -0.085 to -0.084 | [-0.158, -0.011] | ≤0.049 |
| | **Modularity** | 0.088 | [0.020, 0.157] | 0.042 |
| **Fronto-amygdala circuit (R)** | **Topological robustness, Topological stability, Global efficiency** | -0.105 to -0.082 | [-0.171, -0.013] | ≤0.048 |
| | **Modularity** | 0.096 | [0.027, 0.165] | 0.032 |
| **Fronto-basal ganglia-thalamic circuit (bilateral)** | **Global efficiency** | -0.113 to -0.105 | [-0.179, -0.040] | ≤0.016 |
| **Fronto-basal ganglia-thalamic circuit (R)** | **Topological robustness, topological stability, global clustering** | -0.081 to -0.077 | [-0.153, -0.008] | ≤0.049 |
| | **Modularity, fragility** | 0.082 to 0.087 | [0.010, 0.156] | ≤0.049 |
| **Fronto-striatal-thalamic circuit (bilateral)** | **Global efficiency** | -0.113 to -0.105 | [-0.179, -0.040] | ≤0.017 |
| **Central executive (R)** | **Topological robustness, topological stability, global efficiency** | -0.099 to -0.088 | [-0.166, -0.019] | ≤0.030 |
| | **Modularity** | 0.108 | [0.039, 0.177] | 0.021 |
| **Fronto-basal ganglia circuit (bilateral)** | **Global efficiency** | -0.112 to -0.107 | [-0.177, -0.042] | ≤0.013 |
| **Fronto-striatal** | **Global efficiency** | -0.107 | [-0.172, -0.042] | 0.014 |

| | | | | | |
|---|---|---|---|---|---|
| | **circuit (L)** | | | | |
| | **Fronto-basal ganglia-amygdala circuit (bilateral)** | **Global efficiency** | -0.109 to -0.106 | [-0.174, -0.041] | ≤0.014 |
| | **Somatomotor (bilateral)** | **Segregation** | 0.117 to 0.129 | [0.047, 0.200] | ≤0.010 |
| | **Somatomotor (R)** | **Global clustering** | -0.082 | [-0.152, -0.012] | 0.036 |
| | | **Modularity, fragility** | 0.093 to 0.098 | [0.009, 0.178] | ≤0.043 |
| | **Salience (bilateral)** | **Modularity, fragility** | 0.094 to 0.120 | [0.025, 0.190] | ≤0.027 |
| | **Salience (L)** | **Segregation** | 0.123 | [0.046, 0.199] | 0.008 |
| | **Salience (R)** | **Global clustering** | -0.092 | [-0.162, -0.022] | 0.017 |
| | **Default mode (L)** | **Segregation** | 0.110 | [0.040, 0.179] | 0.021 |
| **Social withdrawal** | **Somatomotor (L)** | **Median connectivity (within network), median connectivity (cross-network)** | -0.081 to -0.074 | [-0.146, -0.008] | ≤0.048 |
| | | **Segregation** | 0.080 | [0.013, 0.148] | 0.048 |
| | **Somatomotor (R)** | **Modularity** | 0.081 | [0.013, 0.148] | 0.045 |

**JUNE 2020 SURVEY (#2) SAMPLE**

| Mental health factor | Network | Properties | Beta | 95th % CI | P-value |
|---|---|---|---|---|---|
| **Anxiety** | **Frontoparietal control (bilateral)** | **Global efficiency, global clustering** | 0.090 to 0.099 | [0.016, 0.173] | ≤0.046 |
| | **Frontoparietal control (L)** | **Median connectivity (within network)** | 0.115 | [0.040, 0.190] | 0.028 |
| | **Basal ganglia (bilateral)** | **Median connectivity (within network)** | 0.092 to 0.121 | [0.016, 0.198] | ≤0.043 |
| | **Central executive (R)** | **Median connectivity (within network), global efficiency, global clustering** | 0.081 to 0.120 | [0.008, 0.195] | ≤0.049 |
| **Internalizing behaviors** | **Cerebellum (L)** | **Segregation** | -0.118 | [-0.199, -0.036] | 0.047 |
| **Preference for solitude** | **Temporoparietal (R)** | **Segregation** | 0.117 | [0.036, 0.198] | 0.046 |

**AUGUST 2020 SURVEY (#3) SAMPLE**

| Mental health factor | Network | Properties | Beta | 95th % CI | P-value |
|---|---|---|---|---|---|
| **Anxiety** | **Default mode (R)** | **Median connectivity (within network)** | 0.120 | [0.039, 0.201] | 0.034 |
| | **Reward (R)** | **Median connectivity (within network)** | 0.117 | [0.039, 0.196] | 0.036 |
| **Attention problems** | **Basal ganglia (L)** | **Median connectivity (within network)** | 0.135 | [0.052, 0.217] | 0.007 |

| | | | | | |
|---|---|---|---|---|---|
| | **Limbic (L)** | **Modularity** | -0.125 | [-0.208, -0.042] | 0.033 |
| **Preference for solitude** | **Salience (R)** | **Median connectivity (within network)** | -0.127 | [-0.224, -0.031] | 0.015 |
| | | **Modularity, fragility** | 0.132 to 0.143 | [0.049, 0.227] | ≤0.010 |
| | **Somatomotor (L)** | **Segregation** | 0.124 | [0.040, 0.207] | 0.038 |
| **Social withdrawal** | **Fronto-basal ganglia-thalamic circuit (R)** | **Fragility** | 0.114 | [0.035, 0.193] | 0.049 |
| **OCTOBER 2020 SURVEY (#4) SAMPLE** | | | | | |
| **Mental health factor** | **Network** | **Properties** | **Beta** | **95th % CI** | **P-value** |
| **Anxiety** | **Amygdala-thalamic circuit (L)** | **Modularity** | -0.146 | [-0.245, -0.048] | 0.036 |
| | **Hippocampus (R)** | **Topological stability** | 0.197 | [0.081, 0.312] | 0.005 |
| **Attention problems** | **Fronto-striatal circuit (R)** | **Fragility** | 0.142 | [0.048, 0.235] | 0.031 |
| **DECEMBER 2020 SURVEY (#5) SAMPLE** | | | | | |
| **Mental health factor** | **Network** | **Properties** | **Beta** | **95th % CI** | **P-value** |
| **Depression** | **Hippocampus (R)** | **Topological stability** | 0.179 | [0.040, 0.319] | 0.040 |
| **Anxiety** | **Hippocampus (R)** | **Topological robustness, topological stability** | 0.226 to 0.231 | [0.090, 0.367] | ≤0.006 |
| **Attention problems** | **Dorsal attention (L)** | **Median connectivity (cross-network)** | 0.185 | [0.074, 0.296] | 0.012 |
| **MARCH 2021 SURVEY (#6) SAMPLE** | | | | | |
| **Mental health factor** | **Network** | **Properties** | **Beta** | **95th % CI** | **P-value** |
| **Anxiety** | **Cerebellum (L)** | **Global clustering** | -0.173 | [-0.303, -0.043] | 0.047 |
| | **Temporoparietal (R)** | **Segregation** | 0.317 | [0.187, 0.448] | <0.001 |
| **Attention problems** | **Fronto-striatal circuit (R)** | **Fragility** | 0.220 | [0.092, 0.348] | 0.008 |
| **MAY 2021 SURVEY (#7) SAMPLE** | | | | | |
| **Mental health factor** | **Network** | **Properties** | **Beta** | **95th % CI** | **P-value** |
| **Anxiety** | **Temporoparietal (R)** | **Segregation** | 0.344 | [0.215, 0.472] | <0.001 |
| **Externalizing behaviors** | **Social (L)** | **Global clustering** | 0.142 | [0.017, 0.268] | 0.046 |
| | **Temporoparietal (L)** | **Fragility** | -0.177 | [-0.307, -0.047] | 0.045 |

**Table S8.** Statistics of models testing predictive relationships between pre-pandemic mental health/behavioral problems and topological properties of individual networks in Cohort B (Path B of predictive models). All p-values have been adjusted for the False Discovery Rate. CI: Confidence interval.

| **MAY 2020 SURVEY (#1) SAMPLE** | | | | | |
|---|---|---|---|---|---|
| **Mental health factor** | **Network** | **Properties** | **Beta** | **95th % CI** | **P-value** |
| **Depression** | **Limbic (bilateral)** | **Median connectivity (cross-network)** | 0.080 to 0.108 | [0.026, 0.162] | ≤0.038 |
| | **Limbic (L)** | **Median connectivity (within network)** | 0.079 | [0.024, 0.133] | 0.023 |
| | **Default mode (L)** | **Median connectivity (cross-network)** | 0.078 | [0.024, 0.133] | 0.047 |
| **Anxiety** | **Basal ganglia (bilateral)** | **Median connectivity (cross-network)** | 0.067 to 0.099 | [0.014, 0.153] | ≤0.003 |
| | **Basal ganglia (R)** | **Median connectivity (within network), topological robustness, topological stability, global efficiency, global clustering** | 0.058 to 0.078 | [0.005, 0.132] | ≤0.046 |
| | **Limbic (R)** | **Median connectivity (cross-network)** | 0.085 | [0.032, 0.138] | 0.018 |
| **Attention problems** | **Social (R)** | **Median connectivity (cross-network)** | 0.081 | [0.027, 0.136] | 0.036 |
| | **Frontoparietal control (L)** | **Fragility** | 0.083 | [0.027, 0.139] | 0.038 |
| **Aggressive behavior** | **Social (R)** | **Median connectivity (cross-network)** | 0.087 | [0.034, 0.141] | 0.014 |
| | **Frontoparietal control (R)** | **Fragility** | 0.083 | [0.028, 0.139] | 0.034 |
| | **Central executive (R)** | **Fragility** | 0.085 | [0.031, 0.139] | 0.020 |
| **Internalizing behaviors** | **Basal ganglia (L)** | **Median connectivity (cross-network)** | 0.082 | [0.027, 0.137] | 0.033 |
| **Externalizing behaviors** | **Limbic (R)** | **Median connectivity (cross-network)** | 0.080 | [0.026, 0.134] | 0.035 |
| **Preference for solitude** | **Somatomotor (R)** | **Modularity, fragility** | 0.079 to 0.088 | [0.025, 0.144] | ≤0.022 |
| **JUNE 2020 SURVEY (#2) SAMPLE** | | | | | |
| **Mental health factor** | **Network** | **Properties** | **Beta** | **95th % CI** | **P-value** |
| **Attention problems** | **Fronto-amygdala circuit (L)** | **Fragility** | 0.087 | [0.032, 0.142] | 0.018 |
| **Preference for solitude** | **Somatomotor (R)** | **Global efficiency** | -0.070 | [-0.122, -0.017] | 0.036 |
| | | **Modularity, fragility** | 0.063 to 0.082 | [0.009, 0.136] | ≤0.045 |

| Mental health factor | Network | Properties | Beta | 95th % CI | P-value |
|---|---|---|---|---|---|
| | **Salience (L)** | **Segregation** | 0.082 | [0.026, 0.137] | 0.040 |
| | **Temporoparietal (R)** | **Segregation** | 0.110 | [0.039, 0.181] | 0.026 |
| **Social withdrawal** | **Temporoparietal (R)** | **Segregation** | 0.090 | [0.029, 0.150] | 0.037 |
| **AUGUST 2020 SURVEY (#3) SAMPLE** | | | | | |
| **Mental health factor** | **Network** | **Properties** | **Beta** | **95th % CI** | **P-value** |
| **Depression** | **Amygdala-thalamic circuit (R)** | **Topological stability** | 0.090 | [0.033, 0.147] | 0.017 |
| | | **Fragility** | -0.070 | [-0.129, -0.012] | 0.047 |
| **Preference for solitude** | **Somatomotor (R)** | **Global efficiency** | -0.080 | [-0.134, -0.025] | 0.026 |
| | | **Modularity** | 0.080 | [0.024, 0.136] | 0.026 |
| | **Dorsal attention (R)** | **Fragility** | 0.084 | [0.027, 0.142] | 0.042 |
| | **Salience (L)** | **Segregation** | 0.083 | [0.026, 0.140] | 0.046 |
| | **Temporoparietal (R)** | **Segregation** | 0.117 | [0.054, 0.180] | 0.003 |
| | **Basal ganglia (L)** | **Segregation** | 0.094 | [0.031, 0.157] | 0.034 |
| **OCTOBER 2020 SURVEY (#4) SAMPLE** | | | | | |
| **Mental health factor** | **Network** | **Properties** | **Beta** | **95th % CI** | **P-value** |
| **Depression** | **Amygdala-thalamic circuit (bilateral)** | **Topological stability** | 0.079 to 0.091 | [0.020, 0.151] | ≤0.046 |
| | **Amygdala-thalamic circuit (L)** | **Topological robustness** | 0.084 | [0.025, 0.143] | 0.046 |
| | **Amygdala-thalamic circuit (R)** | **Fragility** | -0.091 | [-0.152, -0.030] | 0.013 |
| **Anxiety** | **Somatomotor (L)** | **Median connectivity (within network)** | -0.076 | [-0.131, -0.021] | 0.035 |
| **Attention problems** | **Somatomotor (R)** | **Global efficiency** | -0.072 | [-0.130, -0.014] | 0.048 |
| | | **Modularity** | 0.070 | [0.011, 0.128] | 0.049 |
| | **Fronto-amygdala circuit (L)** | **Fragility** | 0.094 | [0.035, 0.153] | 0.018 |
| | **Temporoparietal (L)** | **Segregation** | 0.100 | [0.041, 0.159] | 0.010 |
| **Preference for solitude** | **Frontoparietal control (L)** | **Topological robustness, topological stability, global efficiency,** | -0.069 to -0.067 | [-0.125, -0.010] | ≤0.043 |
| | | **Modularity, fragility** | 0.076 | [0.017, 0.134] | 0.043 |
| | **Somatomotor (bilateral)** | **Global clustering** | -0.088 to -0.072 | [-0.144, -0.015] | ≤0.025 |
| | | **Modularity, segregation** | 0.067 to 0.085 | [0.011, 0.142] | ≤0.029 |
| | **Somatomotor (R)** | **Fragility** | 0.070 | [0.012, 0.129] | 0.027 |

| | | | | | |
|---|---|---|---|---|---|
| | **Salience (L)** | **Segregation** | 0.095 | [0.038, 0.153] | 0.012 |
| | **Social (L)** | **Segregation** | 0.087 | [0.030, 0.145] | 0.029 |
| | **Temporoparietal (R)** | **Segregation** | 0.120 | [0.060, 0.181] | 0.001 |
| | **Basal ganglia (L)** | **Segregation** | 0.107 | [0.048, 0.166] | 0.004 |
| **Social withdrawal** | **Prefrontal cortex (R)** | **Median connectivity (within network), global efficiency, global clustering, topological stability** | -0.085 to -0.065 | [-0.140, -0.008] | ≤0.038 |
| | | **Modularity** | 0.070 | [0.015, 0.126] | 0.022 |
| | **Central executive (R)** | **Topological robustness, topological stability, global efficiency, global clustering** | -0.080 to -0.071 | [-0.136, -0.016] | ≤0.031 |
| | | **Modularity** | 0.065 | [0.009, 0.122] | 0.038 |
| | **Fronto-thalamic circuit (R)** | **Global efficiency, global clustering, topological stability** | -0.071 to -0.067 | [-0.126, -0.013] | ≤0.038 |
| | **Fronto-amygdala circuit (R)** | **Global efficiency, global clustering, topological stability** | -0.076 to -0.073 | [-0.131, -0.019] | ≤0.020 |
| | | **Modularity** | 0.061 | [0.006, 0.116] | 0.049 |
| | **Fronto-basal ganglia circuit (R)** | **Global efficiency, global clustering, topological stability** | -0.073 to -0.069 | [-0.129, -0.014] | ≤0.036 |
| | **Fronto-striatal circuit (R)** | **Global efficiency, global clustering, topological stability** | -0.074 to -0.069 | [-0.129, -0.015] | ≤0.034 |
| | **Fronto-basal ganglia-amygdala circuit (R)** | **Global efficiency, global clustering, topological stability** | -0.073 to -0.068 | [-0.128, -0.014] | ≤0.039 |
| | **Somatomotor (R)** | **Global clustering** | -0.075 | [-0.131, -0.018] | 0.019 |
| | | **Modularity** | 0.076 | [0.019, 0.133] | 0.019 |
| | **Limbic (R)** | **Topological stability** | -0.078 | [-0.135, -0.021] | 0.030 |
| | **Temporoparietal (R)** | **Segregation** | 0.116 | [0.058, 0.174] | <0.001 |
| **DECEMBER 2020 SURVEY (#5) SAMPLE** | | | | | |
| **Mental health factor** | **Network** | **Properties** | **Beta** | **95th % CI** | **P-value** |
| **Depression** | **Default mode (L)** | **Median connectivity (within network), median connectivity (cross-network), global clustering** | 0.085 to 0.100 | [0.022, 0.164] | ≤0.021 |
| | **Thalamus** | **Median connectivity (within network), median connectivity (cross-network),** | 0.072 to 0.097 | [0.010, 0.161] | ≤0.039 |

| | | | | | |
|---|---|---|---|---|---|
| | | **topological robustness, topological stability, global clustering** | | | |
| | **Frontoparietal control (L)** | **Median connectivity (cross-network)** | 0.100 | [0.036, 0.164] | 0.021 |
| | **Reward (L)** | **Median connectivity (cross-network)** | 0.089 | [0.028, 0.151] | 0.045 |
| | **Amygdala-thalamic circuit (R)** | **Topological robustness, topological stability** | 0.104 to 0.111 | [0.040, 0.175] | ≤0.008 |
| **Anxiety** | **Amygdala-thalamic circuit (R)** | **Topological robustness, topological stability** | 0.087 to 0.091 | [0.024, 0.153] | 0.033 |
| **Aggressive behavior** | **Default mode (bilateral)** | **Median connectivity (cross-network)** | 0.091 to 0.105 | [0.028, 0.169] | 0.050 |
| | **Default mode (L)** | **Median connectivity (within network)** | 0.090 | [0.026, 0.154] | 0.029 |
| | **Fronto-thalamic circuit (bilateral)** | **Median connectivity (cross-network)** | 0.094 to 0.102 | [0.032, 0.165] | ≤0.029 |
| | **Fronto-thalamic circuit (L)** | **Median connectivity (within network)** | 0.093 | [0.029, 0.156] | 0.022 |
| | **Fronto-amygdala circuit (bilateral)** | **Median connectivity (cross-network)** | 0.090 to 0.100 | [0.028, 0.163] | ≤0.047 |
| | **Fronto-amygdala circuit (L)** | **Median connectivity (within network)** | 0.085 | [0.022, 0.148] | 0.042 |
| | **Fronto-basal ganglia circuit (L)** | **Median connectivity (within network), median connectivity (cross-network)** | 0.093 to 0.099 | [0.030, 0.161] | ≤0.019 |
| | **Fronto-striatal circuit (L)** | **Median connectivity (within network), median connectivity (cross-network)** | 0.090 to 0.099 | [0.027, 0.161] | ≤0.026 |
| | **Fronto-basal ganglia-thalamic circuit (bilateral)** | **Median connectivity (cross-network)** | 0.090 to 0.097 | [0.028, 0.159] | ≤0.047 |
| | **Fronto-basal ganglia-thalamic circuit (L)** | **Median connectivity (within network)** | 0.095 | [0.032, 0.159] | 0.017 |
| | **Fronto-striatal-thalamic circuit (bilateral)** | **Median connectivity (cross-network)** | 0.091 to 0.098 | [0.029, 0.161] | ≤0.041 |
| | **Fronto-striatal-thalamic circuit (L)** | **Median connectivity (within network)** | 0.093 | [0.029, 0.156] | 0.021 |
| | **Fronto-basal ganglia-amygdala circuit (L)** | **Median connectivity (within network), median connectivity (cross-network)** | 0.094 to 0.100 | [0.031, 0.162] | ≤0.018 |
| | **Reward (bilateral)** | **Median connectivity (cross-network)** | 0.088 to 0.093 | [0.027, 0.154] | ≤0.050 |
| | **Social (bilateral)** | **Median connectivity (cross-network)** | 0.102 to 0.124 | [0.039, 0.186] | ≤0.015 |
| | **Limbic (R)** | **Median connectivity** | 0.103 | [0.040, 0.167] | 0.015 |

| Mental health factor | Network | Properties | Beta | 95th % CI | P-value |
|---|---|---|---|---|---|
| | | **(cross-network)** | | | |
| | **Cerebellum (bilateral)** | **Median connectivity (cross-network)** | 0.089 to 0.128 | [0.025, 0.192] | ≤0.037 |
| | **Cerebellum (R)** | **Fragility** | 0.080 | [0.018, 0.143] | 0.039 |
| | **Prefrontal cortex (L)** | **Median connectivity (cross-network)** | 0.103 | [0.040, 0.166] | 0.014 |
| | **Frontoparietal control (R)** | **Fragility** | 0.094 | [0.029, 0.159] | 0.047 |
| **Social withdrawal** | **Somatomotor (R)** | **Modularity** | 0.087 | [0.025, 0.149] | 0.043 |
| | **Cerebellum (R)** | **Modularity** | 0.090 | [0.030, 0.151] | 0.034 |
| | | **MARCH 2021 SURVEY (#6) SAMPLE** | | | |
| **Mental health factor** | **Network** | **Properties** | **Beta** | **95th % CI** | **P-value** |
| **Depression** | **Amygdala-thalamic circuit (R)** | **Topological stability** | 0.094 | [0.031, 0.157] | 0.035 |
| **Anxiety** | **Amygdala-thalamic circuit (R)** | **Topological stability** | 0.087 | [0.026, 0.148] | 0.033 |
| **Attention problems** | **Limbic (L)** | **Median connectivity (cross-network)** | 0.098 | [0.033, 0.162] | 0.030 |
| **Aggressive behavior** | **Limbic (R)** | **Median connectivity (cross-network)** | 0.098 | [0.034, 0.162] | 0.027 |
| **Internalizing behaviors** | **Frontoparietal control (L)** | **Fragility** | 0.093 | [0.031, 0.155] | 0.034 |
| **Preference for solitude** | **Temporoparietal (R)** | **Segregation** | 0.095 | [0.033, 0.157] | 0.026 |
| **Social withdrawal** | **Limbic (L)** | **Modularity** | 0.090 | [0.029, 0.151] | 0.037 |
| | | **MAY 2021 SURVEY (#7) SAMPLE** | | | |
| **Mental health factor** | **Network** | **Properties** | **Beta** | **95th % CI** | **P-value** |
| **Anxiety** | **Salience (bilateral)** | **Modularity** | -0.076 to -0.075 | [-0.137, -0.014] | ≤0.044 |
| | **Salience (R)** | **Median connectivity (within network), global efficiency, global clustering** | 0.067 to 0.092 | [0.008, 0.153] | ≤0.038 |
| | **Social (L)** | **Global clustering** | 0.080 | [0.020, 0.140] | 0.029 |
| | **Amygdala-thalamic circuit (R)** | **Topological stability** | 0.083 | [0.021, 0.145] | 0.042 |
| | **Prefrontal cortex (R)** | **Segregation** | -0.088 | [-0.147, -0.028] | 0.038 |
| **Attention problems** | **Fronto-amygdala circuit (L)** | **Fragility** | 0.112 | [0.049, 0.175] | 0.005 |
| **Aggressive behavior** | **Cerebellum (L)** | **Median connectivity (cross-network)** | 0.089 | [0.027, 0.150] | 0.046 |

| | | | | | |
|---|---|---|---|---|---|
| **Preference for solitude** | **Basal ganglia (L)** | **Segregation** | 0.098 | [0.031, 0.166] | 0.041 |

**Table S9.** Statistics of models testing predictive relationships between pre-pandemic mental health/behavioral problems, mediation by morphometric brain properties, and youth survey-based outcomes during the outbreak in Cohort A (Path D of mediation models). All p-values have been adjusted for the False Discovery Rate. CI: Confidence interval.

| Outcome | Pre-Pandemic Mental Health / Behavioral Problem | Mediation | Statistic | Value | Region | Property | Statistic | Value |
|---|---|---|---|---|---|---|---|---|
| **MAY 2020 (SURVEY #1)** | | | | | | | | |
| **Alone** | **Depression** | **Partial** | **Beta** | 0.146 | **Insula (L)** | **Volume** | **Beta** | 0.143 |
| | | | **95th % Confidence Interval (CI)** | [0.078, 0.214] | | | **95th % CI** | [0.052, 0.235] |
| | | | **P-Value** | <0.001 | | | **P-Value** | 0.006 |
| **Angry** | **Aggressive behavior** | **Partial** | **Beta** | 0.145 | **Superior temporal gyrus (L)** | **Volume** | **Beta** | 0.131 |
| | | | **95th % Confidence Interval (CI)** | [0.075, 0.215] | | | **95th % CI** | [0.036, 0.225] |
| | | | **P-Value** | <0.001 | | | **P-Value** | 0.020 |
| **Angry** | **Externalizing behaviors** | **Partial** | **Beta** | 0.155 | **Superior temporal gyrus (L)** | **Volume** | **Beta** | 0.133 |
| | | | **95th % Confidence Interval (CI)** | [0.082, 0.228] | | | **95th % CI** | [0.040, 0.226] |
| | | | **P-Value** | <0.001 | | | **P-Value** | 0.016 |
| **Angry** | **Externalizing behaviors** | **Partial** | **Beta** | 0.151 | **Cerebellum white matter (R)** | **Volume** | **Beta** | 0.101 |
| | | | **95th % Confidence Interval (CI)** | [0.078, 0.223] | | | **95th % CI** | [0.010, 0.192] |
| | | | **P-Value** | <0.001 | | | **P-Value** | 0.030 |
| **JUNE 2020 (SURVEY #2)** | | | | | | | | |
| **Positive affect** | **Externalizing behaviors** | **Partial** | **Beta** | -0.145 | **Cerebellum cortex (L)** | **Volume** | **Beta** | -0.123 |
| | | | **95th % Confidence Interval (CI)** | [-0.230, -0.059] | | | **95th % CI** | [-0.228, -0.017] |
| | | | **P-Value** | <0.001 | | | **P-Value** | 0.022 |

**Table S10. Statistics of models testing predictive relationships between pre-pandemic mental health/behavioral problems, mediation by morphometric brain properties, and youth survey-based outcomes during the outbreak in Cohort B (Path D of mediation models). All p-values have been adjusted for the False Discovery Rate. CI: confidence interval.**

| Outcome | Pre-Pandemic Mental Health / Behavioral Problem | Mediation | Statistic | Value | Region | Property | Statistic | Value |
|---|---|---|---|---|---|---|---|---|
| **MAY 2020 (SURVEY #1)** | | | | | | | | |
| **Lonely** | **Aggressive behavior** | **Partial** | **Beta** | 0.088 | **Lateral orbitofrontal cortex (R)** | **Volume** | **Beta** | 0.082 |
| | | | **95th % Confidence Interval (CI)** | [0.035, 0.140] | | | **95th % CI** | [0.019, 0.146] |
| | | | **P-Value** | 0.002 | | | **P-Value** | 0.032 |
| **Angry** | **Aggressive behavior** | **Partial** | **Beta** | 0.128 | **Hippocampus (L)** | **Volume** | **Beta** | 0.080 |
| | | | **95th % Confidence Interval (CI)** | [0.074, 0.183] | | | **95th % CI** | [0.008, 0.151] |
| | | | **P-Value** | <0.001 | | | **P-Value** | 0.030 |
| **AUGUST 2020 (SURVEY #3)** | | | | | | | | |
| **Sad** | **Aggressive behavior** | **Partial** | **Beta** | 0.143 | **Superior temporal gyrus (R)** | **Volume** | **Beta** | 0.083 |
| | | | **95th % Confidence Interval (CI)** | [0.088, 0.198] | | | **95th % CI** | [0.015, 0.150] |
| | | | **P-Value** | <0.001 | | | **P-Value** | 0.050 |

**Table S11.** Statistics of models testing moderation of relationships between youth survey-based outcomes during the first 15 months of the pandemic and pre-pandemic mental health/behavioral problems by the topological properties of individual networks across both cohorts. All p-values have been adjusted for the False Discovery Rate. CI: Confidence interval.

| Outcome | Pre-Pandemic Mental Health/Behavioral Problem | Statistic | Value | Brain Network | Property | Interaction Statistic | Interaction Value |
|---|---|---|---|---|---|---|---|
| | | | MAY 2020 SURVEY (#1) | | | | |
| **Mental health** | **Depression, anxiety** | **Beta** | -0.134 to -0.095 | **Somatomotor (L)** | **Median connectivity (within network), median connectivity (cross-network)** | **Beta** | -0.140 to -0.110 |
| | | **95th % CI** | [-0.206, -0.025] | | | **95th % CI** | [-0.212, -0.038] |
| | | **P-Value** | ≤0.040 | | | **P-Value** | ≤0.013 |
| **Perceived stress** | **Depression, social withdrawal** | **Beta** | 0.083 to 0.140 | **Somatomotor (L)** | **Modularity** | **Beta** | -0.084 to -0.083 |
| | | **95th % CI** | [0.030, 0.193] | | | **95th % CI** | [-0.136, -0.031] |
| | | **P-Value** | ≤0.008 | | | **P-Value** | ≤0.015 |
| **Alone** | **Depression** | **Beta** | 0.090 | **Hippocampus (L)** | **Median connectivity (within network)** | **Beta** | 0.106 |
| | | **95th % CI** | [0.023, 0.157] | | | **95th % CI** | [0.038, 0.174] |
| | | **P-Value** | 0.016 | | | **P-Value** | 0.021 |
| | | | JUNE 2020 SURVEY (#2) | | | | |
| **Pandemic-related stress** | **Attention problems; aggressive behavior** | **Beta** | 0.063 to 0.121 | **Frontoparietal control (R)** | **Modularity** | **Beta** | -0.083 to -0.073 |
| | | **95th % CI** | [0.010, 0.175] | | | **95th % CI** | [-0.140, -0.017] |
| | | **P-Value** | ≤0.040 | | | **P-Value** | ≤0.024 |
| **Pandemic-related stress** | **Attention problems** | **Beta** | 0.108 | **Temporoparietal (R)** | **Median connectivity (within network)** | **Beta** | 0.076 |
| | | **95th % CI** | [0.054, 0.162] | | | **95th % CI** | [0.020, 0.132] |
| | | **P-Value** | <0.001 | | | **P-Value** | 0.015 |
| **Pandemic-related stress** | **Attention problems** | **Beta** | 0.114 | **Social (R)** | **Modularity** | **Beta** | -0.069 |
| | | **95th % CI** | [0.060, 0.169] | | | **95th % CI** | [-0.124, -0.013] |
| | | **P-Value** | <0.001 | | | **P-Value** | 0.029 |
| **Pandemic-related stress** | **Attention problems; aggressive behavior** | **Beta** | 0.062 to 0.114 | **Prefrontal cortex (R)** | **Modularity** | **Beta** | -0.111 to -0.078 |
| | | **95th % CI** | [0.009, 0.168] | | | **95th % CI** | [-0.166, -0.025] |
| | | **P-Value** | ≤0.036 | | | **P-Value** | ≤0.008 |
| **Pandemic-related stress** | **Attention problems; aggressive behavior** | **Beta** | 0.057 to 0.113 | **Fronto-thalamic circuit (R)** | **Modularity** | **Beta** | -0.100 to -0.060 |
| | | **95th % CI** | [0.004, 0.167] | | | **95th % CI** | [-0.156, -0.007] |
| | | **P-Value** | ≤0.045 | | | **P-Value** | ≤0.039 |
| **Pandemic-related stress** | **Attention problems; aggressive behavior** | **Beta** | 0.057 to 0.112 | **Fronto-basal ganglia (R)** | **Modularity** | **Beta** | -0.095 to -0.061 |
| | | **95th % CI** | [0.004, 0.166] | | | **95th % CI** | [-0.151, -0.007] |
| | | **P-Value** | ≤0.046 | | | **P-Value** | ≤0.042 |
| **Pandemic-related stress** | **Attention problems;** | **Beta** | 0.058 to 0.113 | **Fronto-striatal circuit (R)** | **Modularity** | **Beta** | -0.094 to -0.060 |

| | | | | | | | |
|---|---|---|---|---|---|---|---|
| | **aggressive behavior** | **95th % CI** | [0.004, 0.166] | | | **95th % CI** | [-0.150, -0.007] |
| | | **P-Value** | ≤0.045 | | | **P-Value** | ≤0.044 |
| **Pandemic-related stress** | **Attention problems; aggressive behavior** | **Beta** | 0.057 to 0.113 | **Fronto-basal ganglia-thalamic circuit (R)** | **Modularity** | **Beta** | -0.092 to -0.057 |
| | | **95th % CI** | [0.004, 0.167] | | | **95th % CI** | [-0.148, -0.004] |
| | | **P-Value** | ≤0.046 | | | **P-Value** | ≤0.044 |
| **Pandemic-related stress** | **Attention problems; aggressive behavior** | **Beta** | 0.057 to 0.113 | **Fronto-striatal-thalamic circuit (R)** | **Modularity** | **Beta** | -0.091 to -0.057 |
| | | **95th % CI** | [0.004, 0.167] | | | **95th % CI** | [-0.147, -0.004] |
| | | **P-Value** | ≤0.046 | | | **P-Value** | ≤0.050 |
| **Pandemic-related stress** | **Attention problems; aggressive behavior** | **Beta** | 0.059 to 0.113 | **Fronto-basal ganglia-amygdala circuit (R)** | **Modularity** | **Beta** | -0.095 to -0.062 |
| | | **95th % CI** | [0.006, 0.167] | | | **95th % CI** | [-0.151, -0.009] |
| | | **P-Value** | ≤0.047 | | | **P-Value** | ≤0.039 |
| **Pandemic-related stress** | **Attention problems; aggressive behavior** | **Beta** | 0.063 to 0.116 | **Central executive (R)** | **Modularity** | **Beta** | -0.082 to -0.068 |
| | | **95th % CI** | [0.009, 0.170 | | | **95th % CI** | [-0.138, -0.013] |
| | | **P-Value** | ≤0.038 | | | **P-Value** | ≤0.025 |
| **Scared** | **Depression** | **Beta** | 0.150 | **Hippocampus (L)** | **Median connectivity (within network)** | **Beta** | 0.102 |
| | | **95th % CI** | [0.081, 0.218] | | | **95th % CI** | [0.035, 0.169] |
| | | **P-Value** | <0.001 | | | **P-Value** | 0.027 |
| **Scared** | **Depression; aggressive behavior** | **Beta** | 0.060 to 0.129 | **Default mode (R)** | **Modularity** | **Beta** | -0.085 to -0.059 |
| | | **95th % CI** | [0.008, 0.182] | | | **95th % CI** | [-0.139, -0.007] |
| | | **P-Value** | ≤0.036 | | | **P-Value** | ≤0.028 |
| **Scared** | **Aggressive behavior** | **Beta** | 0.054 | **Prefrontal cortex (L)** | **Modularity** | **Beta** | -0.066 |
| | | **95th % CI** | [0.001, 0.107] | | | **95th % CI** | [-0.120, -0.011] |
| | | **P-Value** | 0.045 | | | **P-Value** | 0.029 |
| **Scared** | **Aggressive behavior** | **Beta** | 0.053 | **Fronto-basal ganglia-amygdala circuit (L)** | **Modularity** | **Beta** | -0.070 |
| | | **95th % CI** | [0, 0.106] | | | **95th % CI** | [-0.125, -0.015] |
| | | **P-Value** | 0.048 | | | **P-Value** | 0.035 |
| **Scared** | **Attention problems; aggressive behavior** | **Beta** | 0.066 to 0.086 | **Central executive (R)** | **Modularity** | **Beta** | -0.103 to -0.100 |
| | | **95th % CI** | [0.013, 0.140] | | | **95th % CI** | [-0.157, -0.044] |
| | | **P-Value** | ≤0.029 | | | **P-Value** | ≤0.003 |
| **AUGUST 2020 SURVEY (#3)** | | | | | | | |

| | | | | | | | |
|---|---|---|---|---|---|---|---|
| **Sad** | **Aggressive behavior, social withdrawal** | **Beta** | 0.119 to 0.128 | **Somatomotor (L)** | **Modularity** | **Beta** | -0.077 to -0.070 |
| | | **95th % CI** | [0.060, 0.183] | | | **95th % CI** | [-0.132, -0.014] |
| | | **P-Value** | <0.001 | | | **P-Value** | ≤0.030 |
| **Alone** | **Attention problems; aggressive behaviors** | **Beta** | 0.125 to 0.168 | **Temporoparietal (L)** | **Modularity** | **Beta** | -0.116 to -0.089 |
| | | **95th % CI** | [0.069, 0.222] | | | **95th % CI** | [-0.172, -0.034] |
| | | **P-Value** | ≤0.008 | | | **P-Value** | ≤0.008 |

**Table S12. Statistics of models testing moderation of relationships between youth survey-based outcomes during the first 15 months of the pandemic and pre-pandemic mental health/behavioral problems by morphometric brain properties in Cohort A. All p-values have been adjusted for the False Discovery Rate. CI: Confidence interval.**

| Outcome | Pre-Pandemic Mental Health/Behavioral Problem | Statistic | Value | Region | Property | Interaction Statistic | Interaction Value |
|---|---|---|---|---|---|---|---|
| **MAY 2020 SURVEY (#1)** | | | | | | | |
| **Alone** | **Aggressive behavior** | **Beta** | 0.094 | **Frontal pole (R)** | **Volume** | **Beta** | -0.089 |
| | | **95th % Confidence Interval (CI)** | [0.026, 0.162] | | | **95th % CI** | [-0.157, -0.022] |
| | | **P-Value** | 0.007 | | | **P-Value** | 0.028 |
| **JUNE 2020 SURVEY (#2)** | | | | | | | |
| **Pandemic-related stress** | **Internalizing behaviors** | **Beta** | 0.083 | **Nucleus accumbens (R)** | **Volume** | **Beta** | -0.081 |
| | | **95th % CI** | [0.004, 0.162] | | | **95th % CI** | [-0.160, -0.003] |
| | | **P-Value** | 0.041 | | | **P-Value** | 0.043 |
| **OCTOBER 2020 SURVEY (#4)** | | | | | | | |
| **Pandemic-related stress** | **Depression** | **Beta** | 0.130 | **Rostral anterior cingulate cortex (R)** | **Thickness** | **Beta** | -0.170 |
| | | **95th % CI** | [0.031, 0.230] | | | **95th % CI** | [-0.270, -0.069] |
| | | **P-Value** | 0.031 | | | **P-Value** | 0.003 |
| **DECEMBER 2020 SURVEY (#5)** | | | | | | | |
| **Pandemic-related stress** | **Anxiety** | **Beta** | 0.131 | **Superior parietal lobule (R)** | **Volume** | **Beta** | -0.131 |
| | | **95th % CI** | [0.013, 0.249] | | | **95th % CI** | [-0.244, -0.018] |
| | | **P-Value** | 0.029 | | | **P-Value** | 0.037 |

**Table S13.** Statistics of models testing moderation of relationships between youth survey-based outcomes during the first 15 months of the pandemic and pre-pandemic mental health/behavioral problems by morphometric brain properties in Cohort B. All p-values have been adjusted for the False Discovery Rate. CI: Confidence interval.

| Outcome | Pre-Pandemic Mental Health/Behavioral Problem | Statistic | Value | Region | Property | Interaction Statistic | Interaction Value |
|---|---|---|---|---|---|---|---|
| **MAY 2020 SURVEY (#1)** | | | | | | | |
| **Angry** | **Depression; internalizing behaviors** | **Beta** | 0.127 to 0.138 | **Cerebellum white matter (R)** | **Volume** | **Beta** | -0.071 to -0.066 |
| | | **95th % CI** | [0.073, 0.192] | | | **95th % CI** | [-0.124, -0.012] |
| | | **P-Value** | <0.001 | | | **P-Value** | ≤0.017 |
| **Angry** | **Internalizing behaviors** | **Beta** | 0.138 | **Hippocampus (L)** | **Volume** | **Beta** | -0.067 |
| | | **95th % CI** | [0.083, 0.192] | | | **95th % CI** | [-0.121, -0.012] |
| | | **P-Value** | <0.001 | | | **P-Value** | 0.016 |
| **JUNE 2020 SURVEY (#2)** | | | | | | | |
| **Pandemic-related stress** | **Depression, anxiety; internalizing behaviors** | **Beta** | 0.088 to 0.103 | **Nucleus accumbens (R)** | **Volume** | **Beta** | -0.060 to -0.054 |
| | | **95th % CI** | [0.036, 0.156] | | | **95th % CI** | [-0.111, -0.001] |
| | | **P-Value** | ≤0.001 | | | **P-Value** | ≤0.044 |
| **AUGUST 2020 SURVEY (#3)** | | | | | | | |
| **Negative affect** | **Aggressive behavior** | **Beta** | 0.149 | **Entorhinal cortex (L)** | **White matter intensity** | **Beta** | -0.078 |
| | | **95th % CI** | [0.094, 0.204] | | | **95th % CI** | [-0.132, -0.024] |
| | | **P-Value** | <0.001 | | | **P-Value** | 0.013 |
| **Negative affect** | **Externalizing behaviors** | **Beta** | 0.172 | **Medial orbitofrontal cortex (L)** | **White matter intensity** | **Beta** | -0.069 |
| | | **95th % CI** | [0.117, 0.226] | | | **95th % CI** | [-0.123, -0.016] |
| | | **P-Value** | <0.001 | | | **P-Value** | 0.034 |

**Table S14.** Statistics of models testing moderation of relationships between youth survey-based outcomes during the first 15 months of the pandemic and pre-pandemic mental health/behavioral problems by parental engagement in Cohort A. All p-values have been adjusted for the False Discovery Rate. CI: Confidence interval.

| Outcome | Pre-Pandemic Mental Health Problem in Model | Statistic | Value | Interaction Statistic | Interaction Value |
|---|---|---|---|---|---|
| **MAY 2020 SURVEY (#1)** | | | | | |
| **Negative affect** | **Aggressive behavior** | **Beta** | 0.094 | **Beta** | -0.099 |
| | | **95th % Confidence Interval (CI)** | [0.027, 0.161] | **95th % CI** | [-0.170, -0.029] |
| | | **P-Value** | 0.006 | **P-Value** | 0.045 |
| **Angry** | **Aggressive behavior** | **Beta** | 0.128 | **Beta** | -0.106 |
| | | **95th % Confidence Interval (CI)** | [0.058, 0.199] | **95th % CI** | [-0.179, -0.033] |
| | | **P-Value** | <0.001 | **P-Value** | 0.038 |

**Table S15.** Statistics of models testing moderation of relationships between youth survey-based outcomes during the first 15 months of the pandemic and pre-pandemic mental health/behavioral problems by parental engagement in Cohort B. All p-values have been adjusted for the False Discovery Rate. CI: Confidence interval.

| **Outcome** | **Pre-Pandemic Mental Health Problem in Model** | **Statistic** | **Value** | **Interaction Statistic** | **Interaction Value** |
|---|---|---|---|---|---|
| **MAY 2020 SURVEY (#1)** | | | | | |
| **Alone** | **Anxiety** | **Beta** | 0.111 | **Beta** | -0.096 |
| | | **95th % Confidence Interval (CI)** | [0.059, 0.164] | **95th % CI** | [-0.150, -0.042] |
| | | **P-Value** | <0.001 | **P-Value** | 0.004 |

**Table S16:** Distribution of religious beliefs importance in the youth daily life in the entire cohort (n=2641).

| | |
|---|---|
| Not at all important | 609 (23.1%) |
| Not very important | 458 (17.3%) |
| Somewhat important | 791 (30.0%) |
| Very much important | 701 (26.5%) |
| Missing | 82 (3.1%) |

**Table S17.** Statistics of models testing associations between youth spirituality/religiosity and pandemic outcomes after adjustments for pre-pandemic mental health and behavioral issues, across cohorts. All p-values have been adjusted for the False Discovery Rate. CI: Confidence interval.

| Outcome | Spirituality/Religiosity Statistic | Value |
|---|---|---|
| MAY 2020 SURVEY (#1) | | |
| **Negative affect** | **Beta** | -0.154 to -0.080 |
| | **95th % Confidence Interval** | [-0.225, -0.026] |
| | **P-Value** | ≤0.004 |
| **Sad** | **Beta** | -0.197 to -0.089 |
| | **95th % Confidence Interval** | [-0.271, -0.034] |
| | **P-Value** | ≤0.002 |
| **Perceived stress** | **Beta** | -0.109 to -0.061 |
| | **95th % Confidence Interval** | [-0.182, -0.007] |
| | **P-Value** | ≤0.028 |
| **Alone** | **Beta** | -0.137 to -0.089 |
| | **95th % Confidence Interval** | [-0.210, -0.034] |
| | **P-Value** | ≤0.002 |
| **Angry** | **Beta** | -0.116 to -0.065 |
| | **95th % Confidence Interval** | [-0.191, -0.008] |
| | **P-Value** | ≤0.026 |
| AUGUST 2020 SURVEY (#3) | | |
| **Negative affect** | **Beta** | -0.105 to -0.064 |
| | **95th % Confidence Interval** | [-0.189, -0.010] |
| | **P-Value** | ≤0.028 |
| **Lonely** | **Beta** | -0.137 to -0.085 |
| | **95th % Confidence Interval** | [-0.221, -0.030] |
| | **P-Value** | ≤0.003 |
| OCTOBER 2020 SURVEY (#4) | | |
| **Positive affect** | **Beta** | 0.114 to 0.120 |
| | **95th % Confidence Interval** | [0.014, 0.220] |
| | **P-Value** | ≤0.025 |
| **Perceived stress** | **Beta** | -0.137 to -0.082 |
| | **95th % Confidence Interval** | [-0.236, -0.025] |
| | **P-Value** | ≤0.010 |
